# Genome-wide association study of dry eye disease reveals shared heritability with systemic comorbidities

**DOI:** 10.1101/2025.03.18.25324218

**Authors:** Bryan R. Gorman, Jaxon J. Huang, Peter B. Barr, Christopher W. Halladay, Cari L. Nealon, Chris Chatzinakos, Michael Francis, Chen Jiang, Million Veteran Program, Paul B. Greenberg, Wen-Chih Wu, Saiju Pyarajan, Hélène Choquet, Tim B. Bigdeli, Sudha K. Iyengar, Neal S. Peachey, Anat Galor

## Abstract

Dry eye disease (DED) affects up to 25% of the adult population, with chronic symptoms of pain and dryness often negatively impacting quality of life. The genetic architecture of DED is largely unknown. Here, we develop and validate an algorithm for DED in the Million Veteran Program using a combination of diagnosis codes and prescription records, resulting in 132,657 cases and 352,201 controls. In a multi-ancestry genome-wide association study, we identify ten significant loci in nine susceptibility regions with largely consistent effects across ancestries, including loci linked to synapse maintenance (*EPHA5*, *GRIA1*, *SYNGAP1*) and autoimmunity (*BLK*). Phenome-wide scans for genetic pleiotropy indicate substantial genetic correlations of DED with comorbidities, including fibromyalgia, post-traumatic stress disorder, and Sjögren’s disease. Finally, applying genomic structural equation modeling, we derive a latent factor underlying DED and other chronic pain traits which accounts for 51% of the genetic variance of DED.

## INTRODUCTION

Dry eye disease (DED) is a heterogeneous condition defined by a compilation of patient-reported symptoms and physical signs seen on ocular examination^1^. Symptoms of DED include both vision- and pain-related symptoms, although in the majority of cases, pain-related symptoms predominate, with individuals using terms such as “dryness,” “aching,” “grittiness,” and “tenderness” to describe their pain^2^. Biological manifestations of DED observed as signs on ocular examination also vary and can include decreased tear production, epithelial disruption, and tear instability, in isolation or combination^3^. Ocular surface inflammation and tear hyperosmolarity often accompany tear abnormalities^4^. Adding to the complexity of disease, symptoms and signs of DED are often discordant^5^ and individuals may carry a DED diagnosis based on symptoms, signs, or a combination of both. Given this broad definition, it is not surprising that DED is a prevalent condition in the adult population, with an estimated frequency of 5% to 30% worldwide^6^. We previously demonstrated that DED is common in United States (US) veterans, with a prevalence of 19% in male and 22% in female veterans based on International Classification of Disease (ICD) coding and the use of at least one DED-associated medication^7^.

Several conditions and medications have been linked to DED^8^. Common risk factors for DED include female sex, increasing age, medication use (e.g., antihistamines, antidepressants), and adverse environmental conditions such as low humidity and air pollution^6^. In our previous study of US veterans, post-traumatic stress disorder (PTSD) and depression were strongly associated with a DED diagnosis^7^. Other studies have reported associations between DED and pain conditions, including migraine^8^, fibromyalgia, and pelvic pain^9^. DED is also closely related to auto-immune disorders and is one of the defining symptoms of Sjögrens disease (SjD)^10^.

Despite the complexity of defining DED, consistent associations with systemic comorbidities suggest shared heritable factors. Cross-twin cross-trait correlations of DED were higher in monozygotic twins compared to dizygotic twins, suggesting an underlying genetic contribution^11^. Specifically, DED symptoms were estimated to have a heritability of 29% (95% CI: [18%, 40%])^11^. Twin studies in individuals with chronic pain conditions related to DED (e.g., migraine) also reported higher heritability in monozygotic compared to dizygotic twins^12^. Additionally, findings from the TwinsUK Adult Twin Registry suggested shared genetic factors between chronic widespread pain, pelvic pain, irritable bowel syndrome, and DED^13^.

The heterogeneity of DED presentation and a lack of uniformity in International Classification of Diseases (ICD) coding practices for DED have limited the application of biobank-scale genome-wide association studies (GWAS) to date. A Taiwan Biobank (TWB) analysis^14^ of self-reported DED in 14,185 cases and 25,927 controls reported 11 independent loci with suggestive significance (*P* < 1×10^-5^) though none reaching standard genome-wide significance (*P* < 5×10^-8^). A subsequent study found a genetic correlation of 0.19 between DED and depression in TWB^15^. A South Florida study conducted a GWAS of 329 individuals with neuropathic ocular pain (NOP) symptoms (e.g., burning, wind sensitivity)^16^, and found one single nucleotide polymorphism (SNP), rs140293404, met genome-wide significance, suggesting the possibility of genetic variants predisposing an individual to the development of NOP. Similarly, large-scale GWAS on SjD have identified 22 susceptibility loci^17,18^, most of which are linked to immunity-related genes such as *IRF5*, *STAT4*, and *BLK*.

Missing from the literature is an examination of DED heritability in a US population and its phenotypic and genetic relationship to comorbidities. While previous studies identified correlations between various comorbidities and DED^8,9^, it remains unclear whether there are genetic shared risk factors driving all traits. The Million Veteran Program (MVP)^19^ provides an opportunity to bridge this gap as it links genotypic information from a large cohort of US veterans to their medical records. The goal of this study is to examine the heritability of DED in a US veteran population and examine shared genetic factors with DED-associated comorbidities. Given the prevalence of DED and its negative impact on quality of life, a better understanding of disease contributors, including those shared with non-ocular comorbidities, will help develop precision-based treatment algorithms to individual patients.

## METHODS

### Regulatory approval

MVP provides access to health, genetic, and lifestyle information for US veterans who provided informed consent^19^ (https://www.research.va.gov/mvp/). At the time of this study, MVP included genetic data for 658,582 veterans. Local chart reviews were conducted at the eye clinics of three VA Medical Centers (VAMCs): VA Miami Healthcare System (Miami, FL), VA Northeast Ohio Healthcare System (Cleveland, OH), and Providence VA Medical Center (Providence, RI). This study adhered to the tenets of the Declaration of Helsinki and was approved by the VA Office of Research & Development Central Institutional Review Board (VA CIRB 18-41).

### Case-Control algorithm

We devised a case-control algorithm based on ICD-9/10 codes and prescription records. Inclusion criteria for cases included at least two independent diagnoses of DED on different days, based on ICD-9 codes (370.33, 375.15) or ICD-10 codes (H04.121, H04.122, H04.123, H04.129, H16.221, H16.222, H16.223, H16.229, M35.01) and at least one prescription for a medication used for DED (as listed in the drug database) **(Table S1)**. Individuals were included as a control if they had none of the diagnoses of DED listed above and no prescription for any of the DED medications in **Table S1**. To test the algorithm for validity, it was applied to identify cases and controls at Miami, Providence, and Cleveland VAMCs, followed by manual validation of case-control status through chart review at each site. Once satisfied with the overall multi-site positive predictive value (PPV) and negative predictive value (NPV), the validated algorithm was applied to the entire MVP biobank sample to obtain the population for our study. We identified 491,962 individuals overall, with 134,249 DED cases and 357,713 controls.

### Comorbidities

Comorbidities and medication use were gathered from patient-reported survey data (from Lifestyle and Baseline MVP surveys) and electronic medical record (EMR) searches for Charlson Comorbidity Index (CCI) variables.

### Genetic ancestry

Participants in MVP were classified based on similarity to one of five genetic ancestry clusters using external reference panels^20^: European (EUR), African (AFR), American (AMR), East Asian (EAS), and South Asian (SAS). The SAS ancestry cluster was excluded from GWAS analyses due to the small sample size (<100 cases).

### Association of comorbidities and medication use with DED

A multiple logistic regression model was performed for each comorbidity and medication, with DED as the dependent variable, using age, sex, and genetic ancestry as covariates.

### Associations of polygenic scores (PGSs) with DED

Phenome-wide polygenic score files were obtained from the European Molecular Biology Laboratory’s European Bioinformatics Institute (EMBL-EBI) PGS Catalog^21^. The database of PGS scores was downloaded in September 2022. PGSs using MVP in the training data were excluded. All subjects in MVP were scored across all available PGSs using the BCFtools/score plugin (https://github.com/freeseek/score) of BCFtools^22^ v1.1.7. PGSs were then loaded into the dosage format field of VCFs readable by SAIGE for association testing. To determine the pleiotropy of genetic liability to traits on DED, we performed a scan of association between PGSs and DED case-control status using logistic regression models implemented in SAIGE^23^ v1.3.0, adjusting for sex, age, mean-centered age-squared, and 10 ancestry-specific principal components. The GPT-4 Turbo^24^ large language model (version gpt-4-0125-preview) was used to classify PGS phenotypes into broad systemic disease categories for the purposes of visualization.

### GWAS

MVP samples were genotyped on the MVP 1.0 Axiom array as described previously^25^. Genotype data were pre-phased using SHAPEIT4^26^ and imputed to the TOPMed imputation panel version r2 (N=97,256 sequences) using Minimac4^27^. GWAS analyses were performed on ancestry-stratified subsets in MVP using SAIGE^23^ v1.3.0, adjusting for sex, age, mean-centered age-squared, and 10 ancestry-specific principal components. To ensure accurate effect size estimation, Firth approximation was applied to SNPs with *P* < 0.05. Association scans were performed on well-imputed SNPs (INFO>0.5) using an ancestry-specific minor allele frequency (MAF) cutoff of ≥0.5% and a minimum minor allele count (MAC) cutoff of 20.

### GWAS meta-analysis

We performed inverse variance-weighted fixed-effect meta-analyses of the ancestry-stratified GWAS summary statistics. Each set of summary statistics was converted into GWAS-VCFs^28^ using the BCFtools/munge plug-in (https://github.com/freeseek/score) of BCFtools^22^ v1.17, followed by meta-analysis using the BCFtools/metal plug-in (https://github.com/freeseek/score).

### Clumping

We performed clumping to identify independent loci within broad genome-wide significant susceptibility regions. Reference samples were extracted from the EUR superpopulation of the 1000 Genomes Phase 3. We performed clumping using PLINK 1.9^29^ with an *r*^2^ < 0.2 threshold and a 2 Mb window. We further validated that the independent loci identified using this method had *r*^2^ < 0.2 in all ancestries analyzed (EUR, AFR, AMR, and EAS).

### Pleiotropy analysis

We used LDtrait^30^ to perform a scan for pleiotropic associations with each DED locus reported in the GWAS Catalog^31^. Associations with *P* < 5×10^−8^, linkage disequilibrium (LD) *r^2^*> 0.1, and located within 500 kilobases of the DED locus were included. The GWAS Catalog data were retrieved in February 2025.

### Replication analysis

Replication was performed in the multiethnic Genetic Epidemiology Research on Adult Health and Aging (GERA) cohort. The GERA cohort contains genome-wide genotype, clinical, and demographic data of over 110,000 adult members of the Kaiser Permanente Northern California integrated healthcare system^32,33^. GERA samples were genotyped at over 665,000 genetic markers on four ethnic-specific Affymetrix Axiom arrays optimized for European, Latino, East Asian, and African American individuals^34,35^. Imputation was performed by array with Minimac3 v2.0.1^27^, using either the Haplotype Reference Consortium (HRC) reference panel^36^ or the 1000 Genomes Project Phase III reference panel^37^. In GERA, two phenotypes were defined: a *narrow* phenotype (based on DED medication prescription and/or procedure records as listed in **Table S2**), which was most similar to the phenotype used in MVP; and a *broad* phenotype (at least 2 DED diagnoses based on ICD-10 code H04.12 or ICD-9 code 375.15, but no required medication prescription or procedure records). Controls had no DED diagnoses. In GERA, while 3,317 DED cases and 54,516 controls were identified using the *narrow* definition, 16,025 DED cases and 54,818 controls were identified using the *broad* definition. Statistical power was calculated using the Genetic Association Study (GAS) Power Calculator^38^.

### Linkage Disequilibrium Score Regression (LDSC)

Heritability for DED and pairwise genetic correlations (*r_G_*) between DED and other traits were computed using LDSC^39^ v1.0.1. Prior to computing *r_G_*, all summary statistics were quality-controlled and alleles were harmonized to the GRCh38 reference genome using the BCFtools/liftover^40^ and BCFtools/munge plug-ins. European-ancestry LD scores provided by the LDSC software were used, which exclude the major histocompatibility complex (MHC) region as standard practice.

### Genomic Structural Equation Modeling (GenomicSEM)

We applied GenomicSEM^41^ to examine the genetic overlap between DED and other related disorders. GenomicSEM estimates the genetic variance-covariance matrix using LDSC from GWAS summary statistics and is robust to sample overlap. To replicate the multivariate analyses of DED and other disorders performed in twin studies, we performed a single-factor confirmatory factor analysis using summary statistics for DED and other pain-related conditions (fibromyalgia, irritable bowel syndrome, multisite chronic pain, and migraine). We evaluated model fit using the model *χ*^2^ statistic, fit indices (Comparative Fit Index, or CFI; Standardized Root Mean Square Residual, or SRMR), and standardized factor loadings, whereby CFI > 0.9, SRMR < 0.05, and factor loadings > 0.5 were indicators that the model fit the data adequately.

## RESULTS

### Algorithm Validation

A summary of the overall experimental design is provided in **Figure 1**. Validation of the DED algorithm at the Miami VA hospital noted a 100% (41/41) PPV and a 96% (49/51) NPV for differentiating DED cases from controls. This algorithm was then independently validated at two other VA sites (Providence and Cleveland), with a combined PPV of 100% (100/100) and NPV of 98% (98/100). Validation was based on the documentation of DED symptoms, signs, or diagnosis in the clinical note. Across all sites, the PPV was 100% and the NPV was 97%.

**Figure 1.**
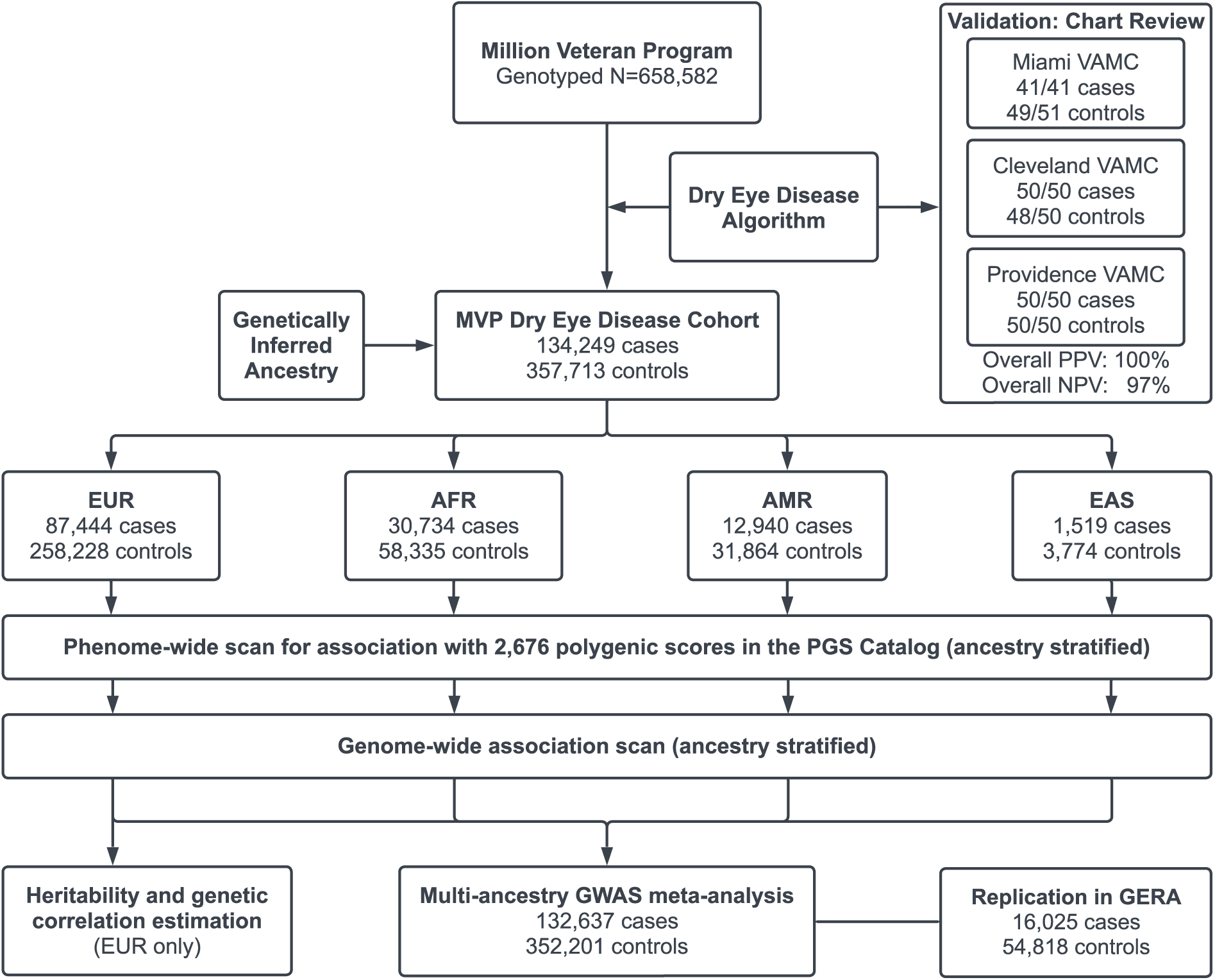
Overall study design. We developed and validated a dry eye disease algorithm based on a combination of diagnosis codes and prescription records and applied it to the Million Veteran Program (MVP) cohort. We inferred the genetic ancestry of participants; four ancestries (EUR, AFR, AMR, and EAS) had sufficient sample size for stratified analytic sets. Polygenic score association scans and GWAS were performed in each ancestry, and GWAS summary statistics were combined in a multi-ancestry meta-analysis. Replication of the 10 genome-wide significant loci was performed in the Genetic Epidemiology Research on Adult Health and Aging (GERA) cohort. The GERA sample size shown corresponds to the GERA broad (ICD code-based) phenotype definition. Heritability, genetic correlation, and genomic structural equation modeling analyses were further performed on the EUR GWAS summary statistics. VAMC: Veterans Affairs Medical Center; EUR: European ancestry; AFR: African ancestry; AMR: American (Hispanic/Latino) ancestry; EAS: East Asian ancestry; PGS Catalog: Polygenic Score Catalog; GWAS: genome-wide association study.

### MVP study population

The total population comprised 658,582 genotyped participants, of which 491,962 individuals (134,249 DED cases and 357,713 controls) met our case-control inclusion criteria **(Figure 1)**. Participants were classified by genetic similarity to five major continental ancestry clusters: European (EUR), African (AFR), American (AMR), East Asian (EAS), and South Asian (SAS)^20^. Four similarity clusters (EUR, AFR, AMR, and EAS) had at least 100 DED cases and thus formed the analytic set for ancestry-stratified genetic analyses (combined 132,657 cases and 352,201 controls). The mean age of the population was 67.8 ± 14.5 years, with the majority having genetically inferred male sex (91.1%) and EUR ancestry (71.8%). Of the total population, 27.1% were classified as a case, meeting the inclusion criteria for a diagnosis of DED. The cases were older, less likely to be male, and more likely to be non-EUR in comparison to the controls **(Table 1)**. Additional comorbidities and medication use in the total population are listed in **Table S3**. Overall, DED cases had poorer health compared to controls, with a higher percentage of comorbidities and medication use in all domains, including neurologic, rheumatologic, and psychiatric.

**Table 1.** Demographics in the study population. Odds ratios and p-values (two-tailed, unadjusted) from multivariate logistic regression models are provided. All models were adjusted for age, sex, and genetic ancestry. For categorical variables, a dash indicates the reference level. N: sample size; DED: dry eye disease; CI: 95% confidence interval; SD: standard deviation; EUR: European ancestry; AFR: African ancestry; AMR: American (Hispanic/Latino) ancestry; EAS: East Asian ancestry; SAS: South Asian ancestry.

| **Demographics** | **Total**<br>**Population**<br>**(n=491,962)** | **Cases**<br>**(DED)**<br>**(n=134,249)** | **Controls**<br>**(No DED)**<br>**(n=357,713)** | **Odds Ratio**<br>**(95% CI)** | **P-value** |
| --- | --- | --- | --- | --- | --- |
| Age, years, mean ± SD | 67.82 ± 14.48 | 72.36 ± 11.09 | 66.13 ± 15.21 | 1.05 (1.04-1.05) | <0.001 |
| Sex, male, n (%) | 441,567 (91.1) | 116,493 (88.6) | 325,074 (92.0) | 2.37 (2.31-2.42) | <0.001 |
| Ancestry, n (%), EUR | 352,392 (71.8) | 87,444 (66.3) | 258,228 (73.8) | - | - |
| AFR | 89,069 (18.4) | 30,734 (23.2) | 58,335 (16.7) | 1.97 (1.94-2.01) | <0.001 |
| AMR | 44,804 (8.7) | 12,940 (9.3) | 31,864 (8.5) | 1.79 (1.75-1.83) | <0.001 |
| EAS | 5,293 (1.0) | 1,519 (1.1) | 3,774 (1.0) | 1.83 (1.71-1.95) | <0.001 |
| SAS | 284 (0.1) | 93 (0.1) | 191 (0.1) | 3.36 (2.59-4.35) | <0.001 |
| Body Mass Index, mean ± SD | 29.67 ± 5.79 | 30.30 ± 5.95 | 29.43 ± 5.72 | 1.04 (1.04-1.04) | <0.001 |
| Smoking status, n (%), Never | 129,539 (26.7) | 35,112 (26.7) | 94,427 (26.7) | - | - |
| Current | 121,867 (25.1) | 30,071 (22.9) | 91,796 (26.0) | 1.07 (1.05-1.09) | <0.001 |
| Prior | 205,942 (42.5) | 64,761 (49.2) | 141,181 (40.0) | 1.16 (1.14-1.18) | <0.001 |

### Phenome-wide polygenic score scan

DED is associated with numerous comorbidities, but it is unclear to what extent these associations are mediated by genetic liability. To comprehensively assess the shared genetic liability between comorbid traits and DED, we performed an unbiased phenome-wide association scan between all polygenic scores in the PGS Catalog and DED case-control status. As presently most polygenic scores have been developed from GWAS in the UK Biobank (UKB) and other European biobanks, we initially focused on the association of PGSs with our EUR DED cohort. Any scores that used MVP as a training set for the base GWAS were excluded. In EUR participants, 574 polygenic scores, representing 195 unique phenotypes mapping to the Experimental Factor Ontology, were significant at a strict Bonferroni cutoff based on the number of scores (*P* < 1.87×10^-5^). The single most predictive PGS of DED case-control status was self-rated health (PGS002218); each standard deviation increase in the PGS was associated with an odds ratio (OR) of 1.14 (95% CI: [1.13, 1.15]). Numerous other PGSs representing genetic liability to diseases across nearly all organ systems were significantly associated with DED **(Figure 2; Table S4)**.

**Figure 2.**
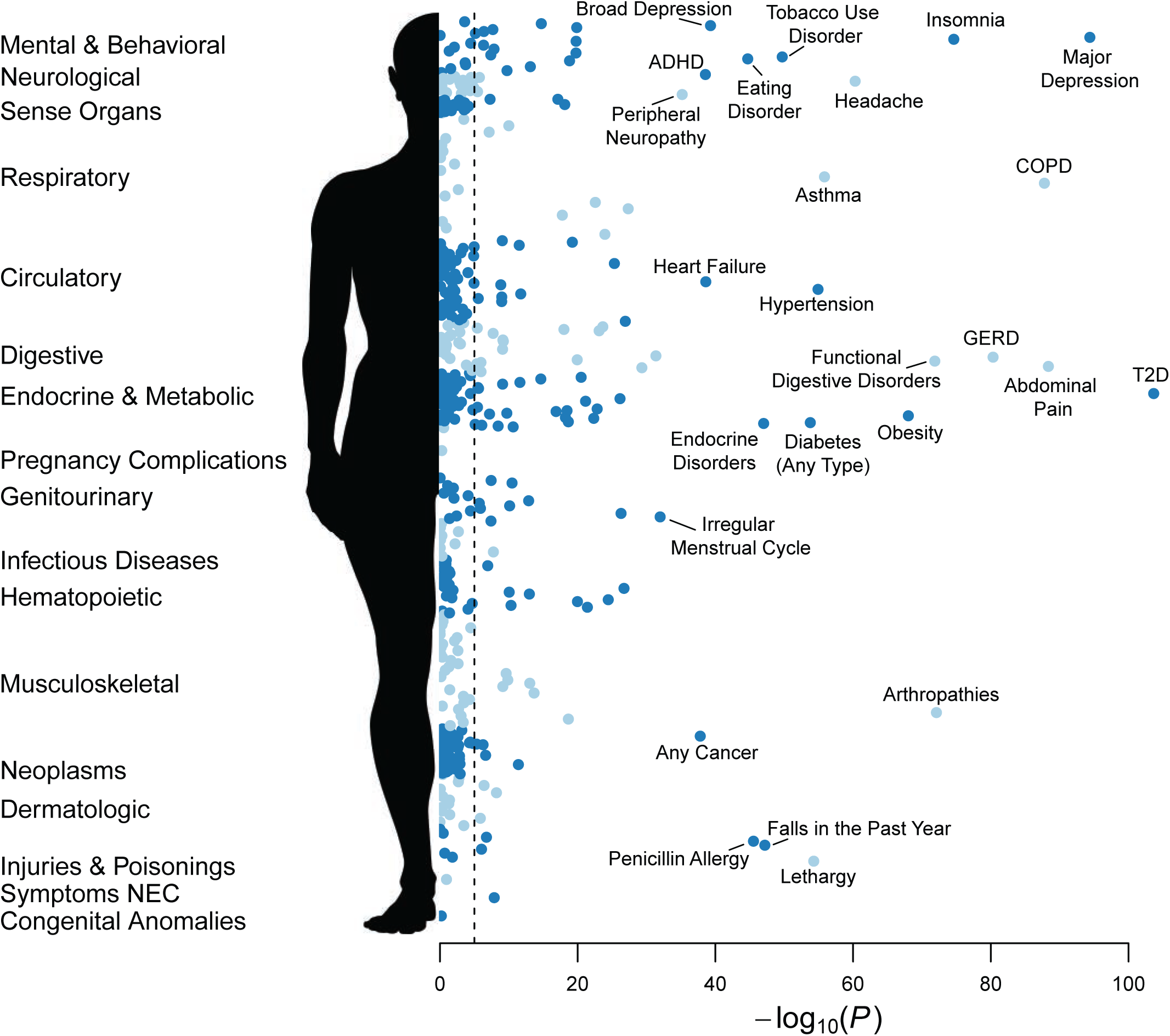
Phenome-wide scan of association between disease-related polygenic scores and dry eye disease (DED) case-control status. Each polygenic score (PGS) in the PGS Catalog was tested for association with DED. Results for European-ancestry participants (87,444 DED cases and 258,228 DED controls) are shown. Full summary statistics are provided in **Table S4**. ADHD: attention deficit hyperactivity disorder; COPD: chronic obstructive pulmonary disorder; GERD: gastro-esophageal reflux disease; T2D: type 2 diabetes.

We then assessed the association of the polygenic scores in other ancestries. In general, the cross-ancestry predictivity of polygenic scores (i.e., transferability) is reduced as a function of genetic distance between the training and target cohorts^42^. The combination of smaller cohort sizes in our non-EUR sets and attenuated cross-ancestry PGS transference greatly reduces statistical power, so a lack of association in a non-EUR ancestry should not be interpreted as evidence of absence. Of the 574 Bonferroni-significant scores in EUR, 179 scores (representing 79 unique traits) were nominally significant (*P* < 0.05) in AFR with directionally consistent effects, 267 scores (106 traits) were nominally significant in AMR, and 13 scores (10 traits) were nominally significant in EAS **(Table S4)**. Association z-scores were highly correlated across ancestries **(Figure S1)**. Six scores (five traits) were Bonferroni-significant in EUR and significant at *P* < 0.05 in AFR, AMR, and EAS, with consistent effect direction: major depressive disorder (MDD) (PGS000907), number of non-cancer illnesses (a survey-based phenotype from UKB; PGS001004), abnormal appetite (PGS002098), neuroticism (PGS002213, PGS001996), and peripheral nerve disorders (PGS002039). Overall, our results are consistent with the genetic liability to DED being shared across ancestries.

### GWAS for DED

We performed a GWAS in each ancestry in MVP, followed by a multi-ancestry meta-analysis. The individual population GWAS and meta-analysis all had genomic inflation factor λ ≤ 1.18 (λ_1000_ ≤ 1.001), indicating minimal systemic inflation of the test statistic **(Figure S2; Table S5)**. In our multi-ancestry meta-analysis, ten independent loci within nine susceptibility regions reached genome-wide significance, all of which were novel associations with respect to DED **(Figure 3; Figure S3; Table 2)**.

**Figure 3.**
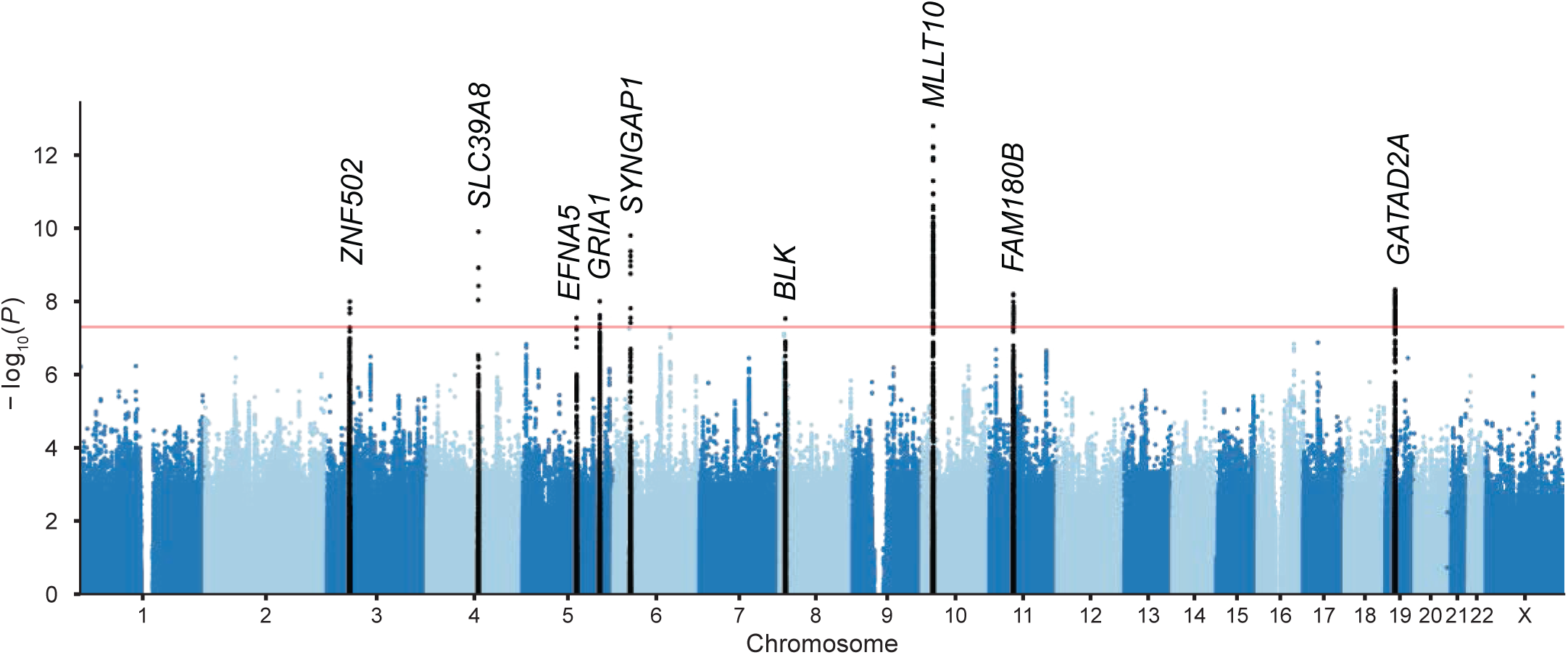
Manhattan plot of GWAS results for dry eye disease. The multi-ancestry meta-anal-ysis of four ancestries represented in the Million Veteran Program is shown (total of 132,637 cases and 352,201 controls). X-axis, chromosome position; Y-axis, −log10 (p-value). The red line denotes the genome-wide significance threshold (*P* < 5×10^−8^). Genome-wide significant loci at each of the nine novel susceptibility regions are highlighted and labeled with the nearest gene.

**Table 2.**
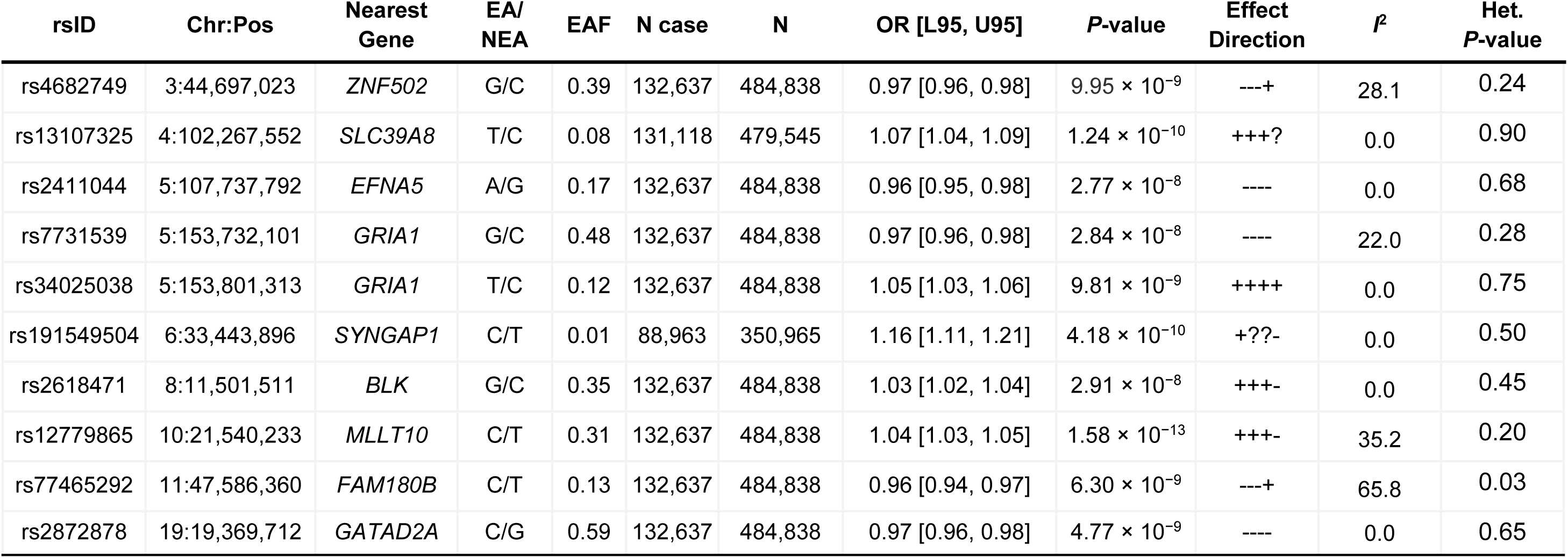
Genome-wide significant loci in the multi-ancestry meta-analysis of dry eye disease (DED). We identified 10 independent loci within 9 susceptibility regions, all of which were novel with respect to DED. Genomic coordinates correspond to the GRCh38 assembly. Ancestry-stratified summary statistics for each locus are provided in **Table S6**. Chr: chromosome; Pos: position; EA: effect allele; NEA: non-effect allele; EAF: effect allele frequency; N: sample size; OR [L95, U95]: odds ratio with lower and upper bounds of the 95% confidence interval; Effect Direction: SNP effect direction in the EUR, AFR, AMR, and EAS cohorts, respectively (“?” indicates the variant did not meet the allele frequency cutoff of 0.1% in that ancestry and was not included in the meta-analysis); I^2^: percentage of variation due to heterogeneity; Het. P-value: p-value of the Cochran’s Q heterogeneity test.

| rsID | Chr:Pos | Nearest Gene | EA/NEA | EAF | N case | N | OR [L95, U95] | P-value | Effect Direction | $I^2$ | Het. P-value |
| --- | --- | --- | --- | --- | --- | --- | --- | --- | --- | --- | --- |
| rs4682749 | 3:44,697,023 | *ZNF502* | G/C | 0.39 | 132,637 | 484,838 | 0.97 [0.96, 0.98] | $9.95 \times 10^{-9}$ | ---+ | 28.1 | 0.24 |
| rs13107325 | 4:102,267,552 | *SLC39A8* | T/C | 0.08 | 131,118 | 479,545 | 1.07 [1.04, 1.09] | $1.24 \times 10^{-10}$ | +++? | 0.0 | 0.90 |
| rs2411044 | 5:107,737,792 | *EFNA5* | A/G | 0.17 | 132,637 | 484,838 | 0.96 [0.95, 0.98] | $2.77 \times 10^{-8}$ | ---- | 0.0 | 0.68 |
| rs7731539 | 5:153,732,101 | *GRIA1* | G/C | 0.48 | 132,637 | 484,838 | 0.97 [0.96, 0.98] | $2.84 \times 10^{-8}$ | ---- | 22.0 | 0.28 |
| rs34025038 | 5:153,801,313 | *GRIA1* | T/C | 0.12 | 132,637 | 484,838 | 1.05 [1.03, 1.06] | $9.81 \times 10^{-9}$ | ++++ | 0.0 | 0.75 |
| rs191549504 | 6:33,443,896 | *SYNGAP1* | C/T | 0.01 | 88,963 | 350,965 | 1.16 [1.11, 1.21] | $4.18 \times 10^{-10}$ | +??- | 0.0 | 0.50 |
| rs2618471 | 8:11,501,511 | *BLK* | G/C | 0.35 | 132,637 | 484,838 | 1.03 [1.02, 1.04] | $2.91 \times 10^{-8}$ | +++- | 0.0 | 0.45 |
| rs12779865 | 10:21,540,233 | *MLLT10* | C/T | 0.31 | 132,637 | 484,838 | 1.04 [1.03, 1.05] | $1.58 \times 10^{-13}$ | +++- | 35.2 | 0.20 |
| rs77465292 | 11:47,586,360 | *FAM180B* | C/T | 0.13 | 132,637 | 484,838 | 0.96 [0.94, 0.97] | $6.30 \times 10^{-9}$ | ---+ | 65.8 | 0.03 |
| rs2872878 | 19:19,369,712 | *GATAD2A* | C/G | 0.59 | 132,637 | 484,838 | 0.97 [0.96, 0.98] | $4.77 \times 10^{-9}$ | ---- | 0.0 | 0.65 |

Of the ten loci, three were genome-wide significant in EUR; otherwise, discovery relied on the added statistical power from combining multiple ancestries **(Table S6)**. Effect sizes were largely consistent across the four ancestry groups; all loci had heterogeneity *P* > 0.05 except for one **(Table 2)**. This locus, rs77465292 (near *FAM180B*), had a stronger effect in AFR (OR = 0.89, 95% CI: [0.84,0.95], *P =* 4.5×10^−4^) than in EUR (OR = 0.96, 95% CI: [0.94,0.97], *P =* 2.3×10^−7^). The three largest populations (EUR, AFR, and AMR) had consistent effect direction across all loci.

All ten loci exhibited pleiotropic associations with numerous other traits, including comorbidities linked to DED **(Table S7)**. For instance, eight loci were linked with body mass index (BMI), eight with insomnia, four with depression or MDD, and two with multi-site chronic pain. One locus, rs2618471 (near *BLK*), had many autoimmune disease pleiotropic associations, including SjD, systemic sclerosis, systemic lupus erythematosus, and rheumatoid arthritis. Interestingly, rs77465292 (near *FAM180B*) was linked with central corneal thickness, corneal resistance factor, and primary open-angle glaucoma (POAG), but the POAG risk allele was protective for DED.

### Replication of genetic results

We sought replication of our GWAS loci in the multiethnic GERA cohort, the largest DED cohort available to us at the time of this study. Demographics of the GERA cohort are found in **Table S8.** Of the ten loci that reached genome-wide significance in MVP, only one (rs12779865) was powered at 80% probability to identify *P* < 0.05 in the GERA broad phenotype and none in the narrow phenotype. Using the broad phenotype, rs12779865 was replicated with OR = 1.03 (95% CI: [1.00,1.06], *P* = 0.026) **(Table S9)**. Overall, seven of the nine loci genotyped in GERA had consistent effect direction with MVP **(Figure S4)**.

### Heritability

We estimated the heritability attributable to additive common variation (SNP-h^2^) using LDSC with the DED EUR summary statistics. The liability scale SNP-h^2^ was 6.0% (standard error 0.37%), assuming a population prevalence of 25%, for a highly significant heritability z-score of 16.1. The LD intercept was 1.04 (s.e. 0.0075), corresponding to an attenuation ratio of 0.15 (s.e. 0.028). Overall, these results indicate that our GWAS is well-calibrated and captures significant heritability, with most of the signal coming from polygenicity as opposed to population stratification.

### Genetic correlation

Based on the results of our PGS association discovery scan, we selected traits that either reached Bonferroni-adjusted significance or were related to significant traits to perform genome-wide genetic correlation analyses. For each trait, we used the largest consortium GWAS where summary statistics were publicly available. Otherwise, we used GWAS from FinnGen^43^, the Pan-UK Biobank project^44^ (Pan-UKB), or, where traits were harmonized across the two biobanks, a meta-analysis of FinnGen and Pan-UKB generated by the FinnGen consortium; details are provided in **Table S10**. For this analysis, we focused on the EUR DED summary statistics. All summary statistics for the other traits were based on European ancestry GWAS, and none included data from MVP. Because these traits were established in the discovery scan, we considered *P* < 0.05 to be statistically significant evidence of genetic correlation.

Of the 54 traits selected for genetic correlation analysis, 52 demonstrated statistically significant genetic correlations, and 40 with *P* < 0.001, consistent with the PGS associations **(Figure 4; Table S11)**. The trait with the highest genetic correlation with DED was fibromyalgia (*r_G_* = 0.69; 95% CI: [0.58,0.81]), followed by multi-site chronic pain (*r_G_* = 0.64, 95% CI: [0.58, 0.70]). Other chronic pain-related traits showed substantial genetic correlation with DED, including gastro-esophageal reflux (*r_G_* = 0.53, 95% CI: [0.45, 0.60]), asthma (*r_G_* = 0.45, 95% CI: [0.38, 0.53]), irritable bowel syndrome (*r_G_* = 0.46, 95% CI: [0.36, 0.56]), and osteoarthritis (*r_G_* = 0.40, 95% CI: [0.33, 0.47]). Notably, a moderate genetic correlation was also found with endometriosis (*r_G_* = 0.32, 95% CI: [0.24, 0.41]), despite the cohort being 90% male, which suggests the association is due to genetic pleiotropy rather than a causal linkage between the traits.

**Figure 4.**
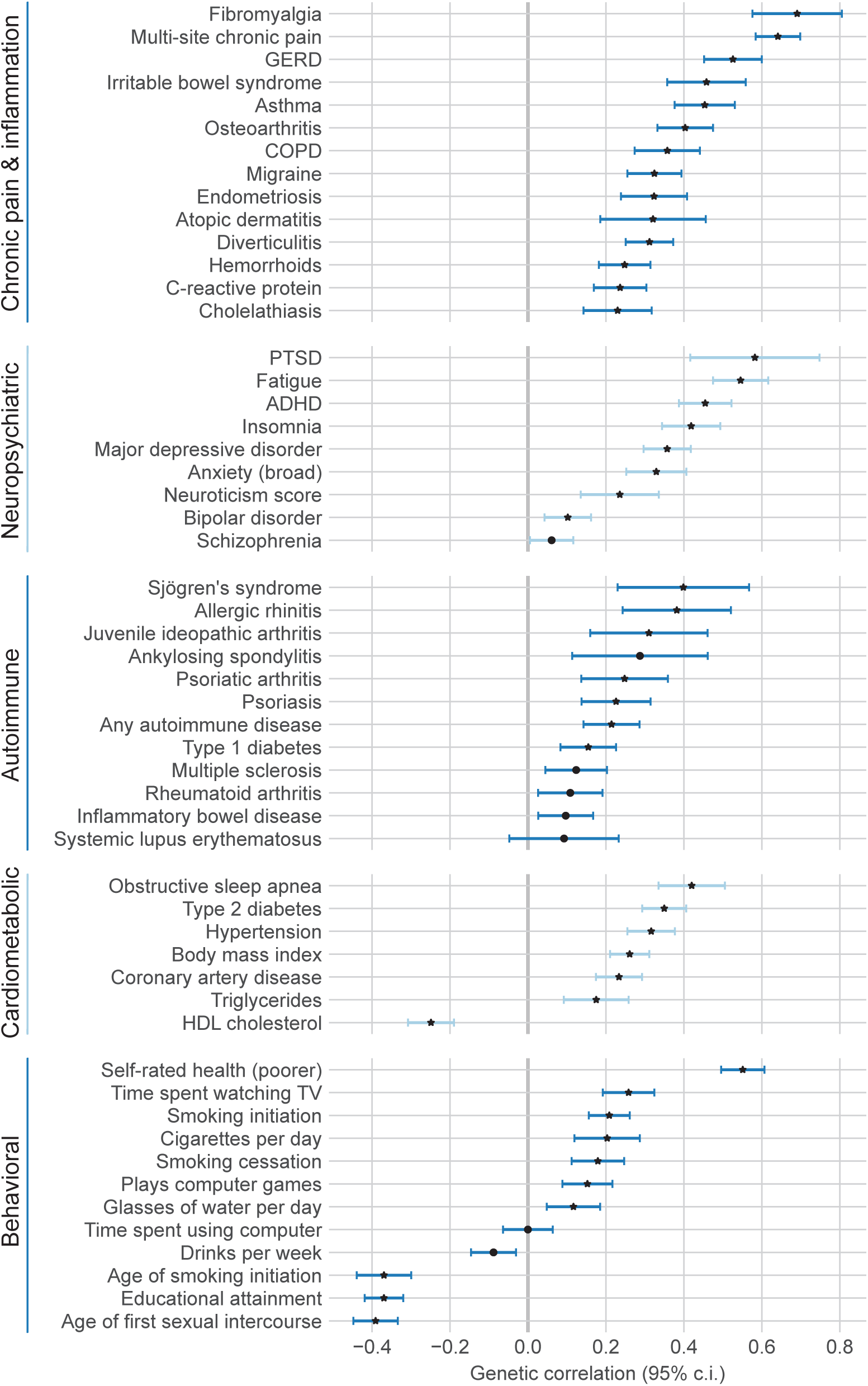
Genetic correlations of selected traits with dry eye disease. Star represents *P* < 0.001 (un-adjusted). GERD: gastro-esophageal reflux disease; COPD: chronic obstructive pulmonary disorder; PTSD: post-traumatic stress disorder; ADHD: attention deficit hyperactivity disorder; HDL cholesterol: high-density lipoprotein cholesterol; 95% c.i.: 95% confidence interval.

Psychiatric and neurological traits also showed substantial genetic correlation with DED. Consistent with our earlier work^7^, the psychiatric trait with the strongest correlation was PTSD (*r_G_* = 0.58, 95% CI: [0.42, 0.75]), followed by attention-deficit/hyperactivity disorder (ADHD) (*r_G_* = 0.45, 95% CI: [0.39, 0.52]), MDD (*r_G_* = 0.36, 95% CI: [0.30, 0.42]), and broadly defined anxiety (*r_G_* = 0.33, 95% CI: [0.25, 0.41]). Bipolar disorder (*r_G_* = 0.10, 95% CI: [0.04, 0.16]) and schizophrenia (*r_G_* = 0.06, 95% CI: [0.01, 0.12]) showed relatively attenuated genetic correlations with DED, consistent with known interrelationships among disorders on the psychosis-affective spectrum^45^. Neurological traits of fatigue (*r_G_* = 0.55, 95% CI: [0.47, 0.62]) and insomnia (*r_G_* = 0.42, 95% CI: [0.34, 0.49]) were also correlated. Finally, neuroticism, a personality trait, was significantly correlated with DED (*r_G_* = 0.24, 95% CI: [0.14, 0.34]).

Autoimmune traits represented another major nexus of genetic correlation with DED. Of autoimmune traits tested, SjD had the highest genetic correlation (*r_G_* = 0.40, 95% CI: [0.23, 0.57]), consistent with dry eyes being a key symptom of SjD. In a sensitivity analysis where individuals with any history of SjD diagnosis were excluded from cases and controls, the genetic correlation with SjD remained virtually the same (*r_G_* = 0.36, 95% CI: [0.19, 0.53]), demonstrating that the genetic correlation with SjD is largely not due to clinical SjD cases. Other autoimmune traits with moderate genetic correlations included allergic rhinitis (*r_G_* = 0.38, 95% CI: [0.24, 0.52]) and juvenile idiopathic arthritis (*r_G_* = 0.31, 95% CI: [0.16, 0.46]). A GWAS based on a diagnosis of any autoimmune disease had *r_G_* = 0.21 (95% CI: [0.14, 0.29]). Notably, these genetic correlations were found despite excluding the MHC region from the analysis, following standard practice.

Cardiometabolic traits also featured prominently. Amongst cardiometabolic traits, sleep apnea had the strongest genetic correlation (*r_G_*= 0.42, 95% CI: [0.33, 0.50]), followed by type 2 diabetes (*r_G_*= 0.35, 95% CI: [0.29, 0.41]), which has reported inflammatory and autoimmune facets. We also found moderate genetic correlations with triglycerides (*r_G_* = 0.18, 95% CI: [0.09, 0.26]) and HDL cholesterol (*r_G_* = -0.25, 95% CI: [-0.31, -0.19]).

Finally, several behavioral traits were genetically correlated with DED. Poor self-rated health was the most correlated behavioral trait (*r_G_*= 0.55, 95% CI: [0.50, 0.61]). Consistent with ADHD, externalizing behavior^46^ was a major theme, featuring genetic correlations with earlier tobacco use (*r_G_*= -0.37, 95% CI: [-0.44, -0.30]), earlier sexual debut (*r_G_* = -0.39, 95% CI: [-0.45, -0.33]), and lower educational attainment (*r_G_*= -0.37, 95% CI: [-0.42, -0.32]). Whereas the PGS for general screen usage (computer or TV) was associated with DED, here we assessed genetic correlation with computer usage and TV usage separately. While time spent watching TV (*r_G_* = 0.26, 95% CI: [0.19, 0.32]) and gaming (*r_G_* = 0.15, 95% CI: [0.09, 0.22]) were correlated, general computer usage was not (*r_G_*= 0.00, 95% CI: [-0.06, 0.06]). Interestingly, alcohol use measured in drinks per week was negatively correlated *r_G_* = -0.09, 95% CI: [-0.15, -0.03]), potentially reflecting the complex genetic relationship between alcohol use and MDD and other psychiatric traits^47^. Finally, we observed a genetic correlation with increased water intake (*r_G_* = 0.12, 95% CI: [0.05, 0.19]), consistent with observational associations^48^.

### GenomicSEM

**Figure 5** presents the results from GenomicSEM analyses of DED, fibromyalgia, irritable bowel syndrome, multisite chronic pain, and migraine. Overall, there were moderate to large genetic correlations across each of the phenotypes **(Figure 5A)**. **Figure 5B** presents a single factor model, which showed good fit to the data (*χ*^2^ = 3.91, degrees of freedom (df) = 5, *P* = 0.56; comparative fit index (CFI) = 0.99; standardized root mean square residual (SRMR) = 0.02). All of the indicators displayed moderate-to-large loadings (i.e., ≥ 0.5) on the single factor, and the residual variance of DED was 0.49. Lastly, the latent “pain” factor showed relatively strong genetic correlations with other autoimmune disorders, psychiatric conditions such as depression or PTSD, and cardiometabolic outcomes, among others **(Figure 5C)**. Compared to the bivariate genetic correlations between DED and each of these phenotypes, the estimates often became stronger and more precise **(Table S12)**.

**Figure 5.**
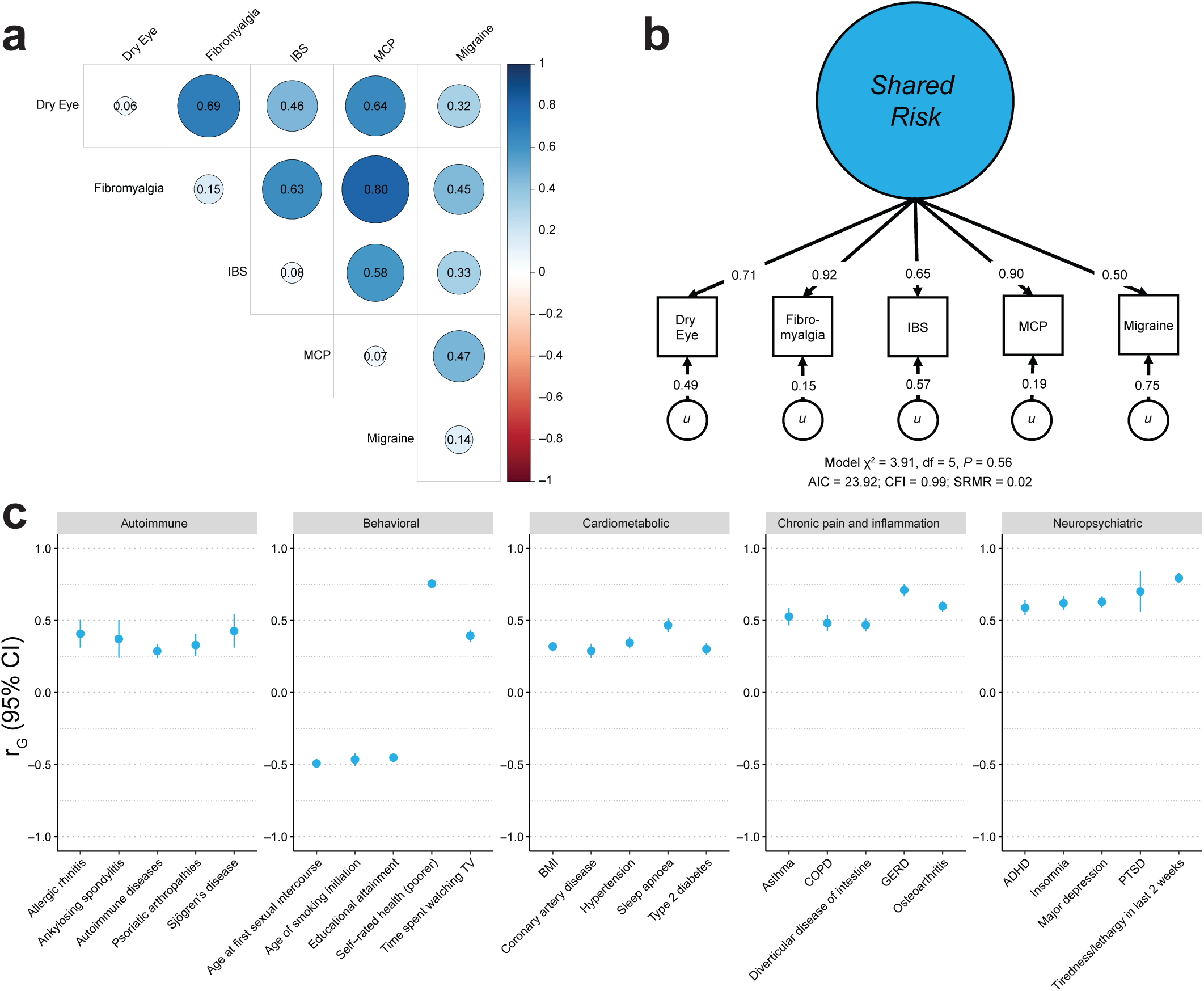
Genomic structural equation modeling of DED identifies shared pain risk. **a)** Heatmap of pairwise genetic correlations between DED and representative chronic pain traits. The SNP heritability (SNP-*h*^2^) is represented along the diagonal. **b)** The genomic SEM model used to derive a latent factor representing the shared genetic risk underlying DED and other chronic pain traits. The arrows pointing from the shared latent factor to each trait represent the factor loading, and the arrows pointing from *u* to each trait represent the residual variance. Model fit indices are provided below the model diagram. **c)** Forest plot of genetic correlations of the latent factor with DED-linked traits of interest. IBS: irritable bowel syndrome; MCP: multisite chronic pain; SEM: structural equation modeling; *u*: residual; df: degrees of freedom; AIC: akaike information criterion; CFI: comparative fit index; SRMR: standardized root mean square residual; r_G_: genetic correlation; 95% CI: 95% confidence interval; BMI: body mass index; COPD: chronic obstructive pulmonary disorder; GERD: gastro-esophageal reflux disease; ADHD: attention defi-cit hyperactivity disorder; PTSD: post-traumatic stress disorder.

## DISCUSSION

This multi-ancestry GWAS meta-analysis of 132,637 DED cases and 484,838 controls in MVP identified ten loci in nine susceptibility regions. To our knowledge, these loci are the first to be associated with DED at the accepted genome-wide significance threshold (*P* < 5×10^−8^). All loci had widespread multi-systemic pleiotropy with other traits, with generally consistent direction of effects with known DED-linked comorbidities. Several loci were within or near genes linked to plausible DED disease mechanisms, such as autoimmunity (*BLK*) and neurotransmitter signaling and synaptic plasticity (*SYNGAP1*, *GRIA1*, *EFNA5*). Notably, all loci had relatively small effect sizes; the largest effect was OR=1.16 at rs191549504 (*SYNGAP1*), a missense mutation with a frequency of just 1% in EUR **(Figure S5)**. These observations are consistent with DED being a highly polygenic trait, with many loci having at most a minor contribution to the liability. Additionally, this illustrates the importance of large sample sizes, such as those provided by MVP, for reliably detecting individual loci associated with DED. In that sense, we found the genetic architecture of DED is more similar to psychiatric traits, such as depression^49^, than other complex ocular disease traits, such as age-related macular degeneration^50^ or glaucoma^51^, which are partially characterized by common loci with large effect sizes. Our replication analysis confirmed the most significant locus from our GWAS (at *MLLT10*) but was underpowered for statistical significance in the remaining loci, confirmation of which will require future multi-biobank analyses.

To better understand genetic relationships between DED and systemic comorbidities, we performed a phenome-wide polygenic score association scan, followed by genome-wide genetic correlation analyses. Our results were largely consistent with known observational associations between DED and various systemic comorbidities, demonstrating that shared genetic risk factors partially underlie these relationships. Notably, DED genetic risk exhibits multisystemic pleiotropy across neuropsychiatric, autoimmune, cardiometabolic, and chronic pain-related disease traits. We further jointly modeled several chronic pain traits using genomicSEM to derive a latent factor representing shared genetics between DED and other chronic pain traits. We found that our latent factor had a loading of 0.7 on DED, explained 51% of the genetic variance of DED, and itself exhibited multisystemic pleiotropy. This suggests that DED may be an indicator of poor health status across multiple systems in the human body.

Our findings help reframe the conversation regarding disease correlations, highlighting caution in imputing causation. Our results suggest that many traits are likely associated with DED through shared underlying etiology, manifested in a high genetic correlation. For instance, digital screen usage has been associated with DED in numerous observational studies and is generally assumed to be causal of DED through altered blinking dynamics or other mechanisms^52^. Yet our analysis suggests that associations with DED and screen usage may be at least partially confounded. DED was associated with the polygenic score for overall screen usage, but breaking it down into types of screen usage, we observed significant genetic correlations with TV usage (*r_G_* = 0.26; 95% CI: [0.19, 0.32]) and gaming (*r_G_*= 0.15; 95% CI: [0.09, 0.22]), but not general computer usage (*r_G_* = 0.00; 95% CI: [-0.06, 0.06]). Notably, the chronic pain latent factor was even more genetically correlated with TV usage (*r_G_* = 0.39; 95% CI: [0.35, 0.44]) and gaming (*r_G_* = 0.21; 95% CI: [0.17, 0.25]) than DED was, implying that underlying factors are more likely to tie together the noted associations than one trait causing the other. These findings have therapeutic implications as current algorithms generally treat such associations as casual, suggesting modulation of screen time, for example, as a treatment for DED, rather than identifying root causes that tie specific screen use and certain DED sub-types together. Future studies are therefore needed to examine genetically informed causal inferences as applied to DED and DED sub-types to develop optimized treatment algorithms for individual patients.

Our work has several limitations. First, our phenotyping algorithm was based on EMR diagnoses and prescriptions, which limits our ability to identify associations with specific DED symptoms and signs. Secondly, our study was performed in a population of military veterans receiving care at the VA, which may limit the generalizability of the results to other populations. Importantly, with respect to DED, which is more prevalent in females, our cohort was predominantly male and therefore our results could potentially reflect male-specific risk factors. Sex-stratified analyses will be the focus of future work. Finally, while our PGS scan and GWAS analyses were performed across all four major continental ancestry groups in MVP and demonstrated generally consistent results across ancestries, our heritability and genetic correlation analyses focused on EUR individuals. This was due to technical limitations of LDSC with respect to genetically admixed populations and a present lack of GWASs across many of the traits of interest in AFR and AMR populations. As these data and new methods become available over time, we plan to update this analysis to be more inclusive.

Despite these limitations, our study adds to the available literature as we successfully developed and validated a DED phenotyping algorithm that can be applied to other EMR-linked biobanks. In addition to identifying the first GWAS loci for DED, we attained a significant overall heritability, allowing us to examine phenome-wide genetic correlations with DED. Our results further motivate future analyses examining genetic correlations with specific signs and symptoms that comprise DED subtypes. Overall, our results provide novel insights into the biological basis of DED and help unify years of observational associations of DED with systemic comorbidities and behavioral traits.

## Data Availability

The full summary level association data from the meta-analysis and individual population association analyses in MVP will be available via the dbGaP study accession number phs001672.

## ACKNOWLEDGEMENTS

This research is based on data from the Million Veteran Program (MVP), Office of Research and Development, Veterans Health Administration, and was supported by MVP000 and the Veterans Affairs Cooperative Studies Program, study no. 575B. This work also received funding from the Department of Veterans Affairs Office of Research and Development grants EPID-006-15S (Dr. Galor), I01 CX002015 (Dr. Galor), I01 BX004893 (Dr. Galor), I01 BX004557 (Dr. Peachey), IK6 BX005233 (Dr. Peachey), I21 RX003883 (Dr. Galor), Department of Defense Gulf War Illness Research Program W81XWH-20-1-0579 (Dr. Galor) and Vision Research Program (VRP) W81XWH-20-1-0820 (Dr. Galor), National Eye Institute U01 EY034686 (Dr. Galor), R01 EY026174 (Dr. Galor), R61 EY032468 (Dr. Galor), NIH Center Core Grant P30EY014801 (University of Miami), P30EY025885 (Cole Eye Institute Core), P30EY011377 (Case Western Reserve University Core), and unrestricted awards from Research to Prevent Blindness to the Departments of Ophthalmology of University of Miami and Cleveland Clinic Lerner College of Medicine of Case Western Reserve University. We are grateful to R. Karlsson Linnér, PhD (Leiden University), and T.T. Mallard, PhD (Massachusetts General Hospital), for sharing their script for plotting PheWAS results, which served as the basis of the figure presented herein. This publication does not represent the views of the Department of Veterans Affairs or the United States Government. We want to acknowledge the participants and investigators of the Million Veteran Program, the Genetic Epidemiology Research on Adult Health and Aging (GERA) cohort, the FinnGen study, the UK Biobank, and the Pan-UKB project. Full consortium acknowledgements for the Million Veteran Program are provided in the Supplementary Information.

No conflicting relationships exist for any author.

## SUPPLEMENTARY CONTENT

**Supplementary Information.** VA Million Veteran Program: Core Acknowledgements

**Figure S1.** Comparison of polygenic score associations across ancestries. Disease-related polygenic risk scores from the PGS Catalog were calculated in MVP and associated with dry eye disease case-control status in each ancestry. For comparison purposes, scores for the same phenotype were de-duplicated by semantic mapping to EFO terms, selecting the score for a harmonized EFO phenotype that was most significant in EUR. Correlation coefficients were calculated over all de-duplicated scores and over those with FDR < 0.05 significance in both ancestries. a) EUR (87,444 cases and 258,228 controls) versus AFR (30,734 cases and 58,335 controls) PGS z-scores. b) Comparison of EUR and AMR (12,940 cases and 31,864 controls) PGS z-scores.

**Figure S2.** GWAS Q-Q plots. Genomic control inflation values (λ) are provided for each GWAS. a) EUR (87,444 cases and 258,228 controls). b) AFR (30,734 cases and 58,335 controls). c) AMR (12,940 cases and 31,864 controls). d) EAS (1,519 cases and 3,774 controls). e) Multi-ancestry meta-analysis (132,637 cases and 352,201 controls).

**Figure S4.** Comparison of effect sizes at GWAS loci between MVP and the GERA replication cohorts. a) MVP multi-ancestry meta-analysis (132,637 cases and 352,201 controls) versus the GERA *broad* phenotype (16,025 cases and 54,818 controls). b) MVP multi-ancestry meta-analysis versus the stricter GERA *narrow* phenotype (3,317 cases and 54,516 controls). Log odds ratios with 95% confidence intervals are shown. One GWAS locus (rs191549504, at *SYNGAP1*) was not genotyped in GERA. The *MLLT10* locus (rs12779865) is labeled.

**Figure S5.** Comparison of effect size and allele frequency for GWAS loci. Scatter plot of odds ratio versus risk allele frequency for risk loci in the multi-ancestry meta-analysis (132,637 cases and 352,201 controls). The blue line denotes 80% power to achieve genome-wide significance. Independent genome-wide significant loci are highlighted in blue and labeled by the nearest gene. Additional independent loci reaching a suggestive significance threshold (P < 1×10^−6^) are shown in gray.

**Table S1.** Dry eye disease medications included as inclusion criteria for cases in MVP, as listed in the VA drug database.

**Table S2.** Dry eye disease medications and procedures included as inclusion criteria for cases in the Genetic Epidemiology Research on Adult Health and Aging (GERA) cohort.

**Table S3.** Association of comorbidities and medications with dry eye disease in MVP. Each row represents a multivariate logistic regression with adjustment for age, sex, and ancestry. Odds ratios (95% confidence intervals) and p-values (two-tailed, adjusted for multiple hypothesis testing) are provided.

**Table S4.** Polygenic score associations with dry eye disease. Polygenic scores from the PGS Catalog were calculated in MVP and associated with dry eye disease case-control status in each ancestry. Beta values for each ancestry represent the log-odds ratio associated with a standard deviation increase in the PGS. Summary statistics are sorted by p-value in the EUR population.

**Table S5.** Genome wide association study (GWAS) inflation factors stratified by ancestry.

**Table S6.** Ancestry-stratified summary statistics for the genome-wide significant dry eye disease loci. NA indicates the locus fell below the minor allele frequency (MAF > 0.1%), minor allele count (MAC > 20), or imputation quality (INFO > 0.5) thresholds within the ancestry and was thus excluded from the GWAS and subsequent multi-ancestry meta-analysis.

**Table S7.** Pleiotropic associations with dry eye disease risk alleles. LDtrait was used to scan for reported GWAS associations at, or in linkage disequilibrium (LD) with, DED risk loci. The 1000 Genomes high-coverage EUR reference panel was used for calculation of R2 and D’ values.

**Table S8.** Genetic Epidemiology Research on Adult Health and Aging cohort demographics.

**Table S9.** Replication of genome-wide association study (GWAS) findings in the Genetic Epidemiology Research on Adult Health and Aging (GERA) cohort. Replication in GERA was analyzed across narrow and broad phenotype definitions. Only one SNP for the broad phenotype, highlighted in bold, was powered to obtain a *P* < 0.05 significance threshold with 80% probability.

**Table S10.** Metadata of genome-wide association studies (GWASs) of selected traits used in genetic correlation and genomic structural equation modeling (genomicSEM) analyses. Sample prevalence: prevalence of cases in the GWAS cohort. Population prevalence: estimated prevalence in the population.

**Table S11.** Genetic correlations of selected traits with dry eye disease. Traits were significant, or related to significant traits, in the phenome-wide polygenic score association scan.

**Table S12.** Genetic correlations of selected traits with the latent factor produced by the genomic structural equation modeling (GenomicSEM) model.

