## Supplementary Information for "Genome-wide association study of dry eye disease reveals shared heritability with systemic comorbidities"

[1]Center for Data and Computational Sciences (C-DACS), VA Boston Healthcare System, 150 S Huntington Avenue, Boston, MA, 02130, USA, [2]Surgical and Research Services, Miami Veterans Administration Medical Center, 1201 NW 16th Street, Miami, FL, 33125, USA, [3]Bascom Palmer Eye Institute, University of Miami, 900 NW 17th Street, Miami, FL, 33136, USA, [4]Research Service, VA New York Harbor Healthcare System, Brooklyn, NY, 11209, USA, [5]Department of Psychiatry and Behavioral Sciences, SUNY Downstate Health Sciences University, Brooklyn, NY, 11203, USA, [6]Institute for Genomics in Health, SUNY Downstate Health Sciences University, Brooklyn, NY, 11203, USA, [7]Epidemiology & Biostatistics, School of Public Health, SUNY Downstate Health Sciences University, Brooklyn, NY, 11203, USA, [8]Center of Innovation in Long Term Services and Supports, Providence VA Medical Center, 830 Chalkstone Avenue, Providence, RI, 02908, USA, [9]Eye Clinic, VA Northeast Ohio Healthcare System, 10701 East Boulevard, Cleveland, OH, 44106, USA, [10]Ophthalmology & Visual Sciences, Case Western Reserve University School of Medicine, Cleveland, OH, 44106, USA, [11]Division of Research, Kaiser Permanente Northern California (KPNC), 4480 Hacienda Drive, Pleasanton, CA, 94588, USA, [12]Ophthalmology Section, Providence VA Medical Center, 830 Chalkstone Avenue, Providence, RI, 02909, USA, [13]Division of Ophthalmology, Alpert Medical School, Brown University, Providence, RI, 02903, USA, [14]Cardiology Section, Medical Service, Providence VA Medical Center, 830 Chalkstone Avenue, Providence, RI, 02908, USA, [15]Department of Health Systems Science, Kaiser Permanente Bernard J. Tyson School of Medicine, Pasadena, CA, 91101, USA, [16]Cleveland Institute for Computational Biology, Case Western Reserve University, Cleveland, OH, 44106, USA, [17]Department of Population and Quantitative Health Sciences, Case Western Reserve University School of Medicine, Cleveland, OH, 44106, USA, [18]Research Service, VA Northeast Ohio Healthcare System, 10701 East Boulevard, Cleveland, OH, 44106, USA, [19]Cole Eye Institute, Cleveland Clinic Foundation, 9500 Euclid Avenue, Cleveland, OH, 44195, USA, [20]Department of Ophthalmology, Cleveland Clinic Lerner College of Medicine of Case Western Reserve University, 9500 Euclid Avenue, Cleveland, OH, 44195, USA, *These authors contributed equally: B.R.G., J.J.H., †These authors jointly supervised this work: S.K.I., N.S.P., A.G., ‡Corresponding authors:

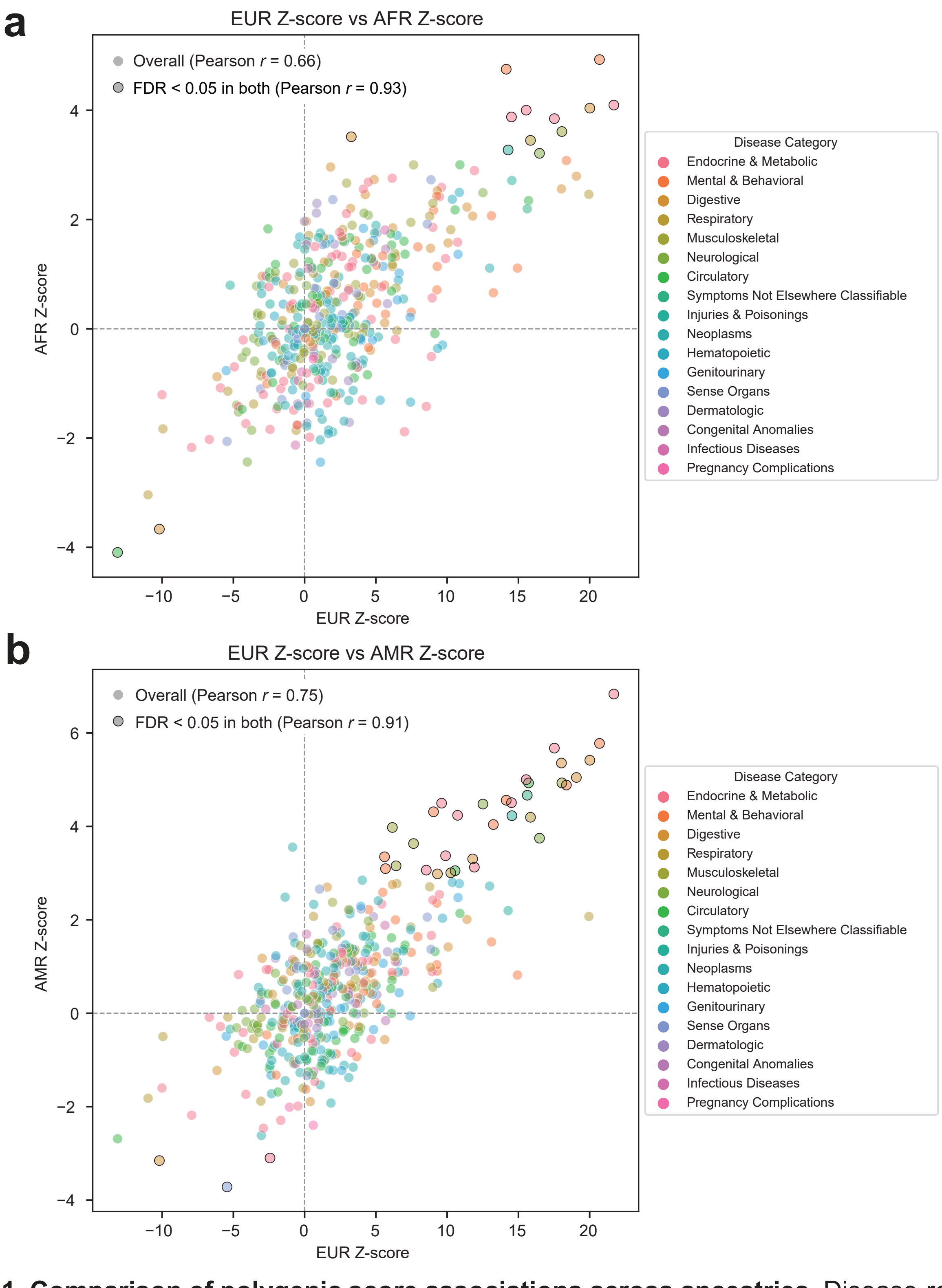


**Figure S1. Comparison of polygenic score associations across ancestries.** Disease-related polygenic risk scores from the PGS Catalog were calculated in MVP and associated with dry eye disease case-control status in each ancestry. For comparison purposes, scores for the same phenotype were de-duplicated by semantic mapping to EFO terms, selecting the score for a harmonized EFO phenotype that was most significant in EUR. Correlation coefficients were calculated over all de-duplicated scores and over those with FDR < 0.05 significance in both ancestries. **a)** EUR (87,444 cases and 258,228 controls) versus AFR (30,734 cases and 58,335 controls) PGS z-scores. **b)** Comparison of EUR and AMR (12,940 cases and 31,864 controls) PGS z-scores.

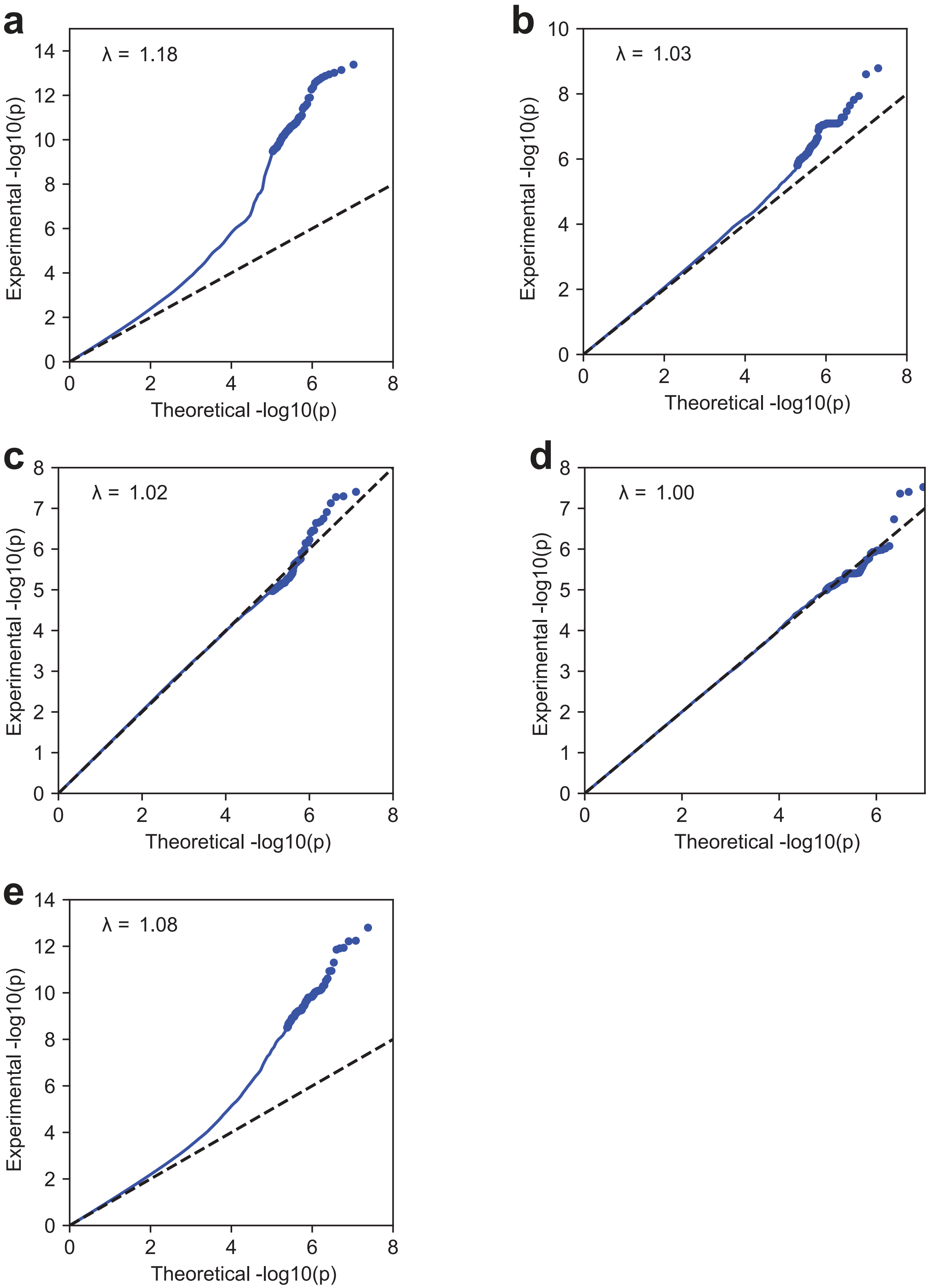


**Figure S2. GWAS Q-Q plots.** Genomic control inflation values (λ) are provided for each GWAS. **a**) EUR (87,444 cases and 258,228 controls). **b**) AFR (30,734 cases and 58,335 controls). **c**) AMR (12,940 cases and 31,864 controls). **d**) EAS (1,519 cases and 3,774 controls). **e**) Multi-ancestry meta-analysis (132,637 cases and 352,201 controls).

**Figure S3. Regional association plots of significant loci from the multi-ancestry meta-analysis.** 1000 Genomes EUR samples were used for the LD reference.

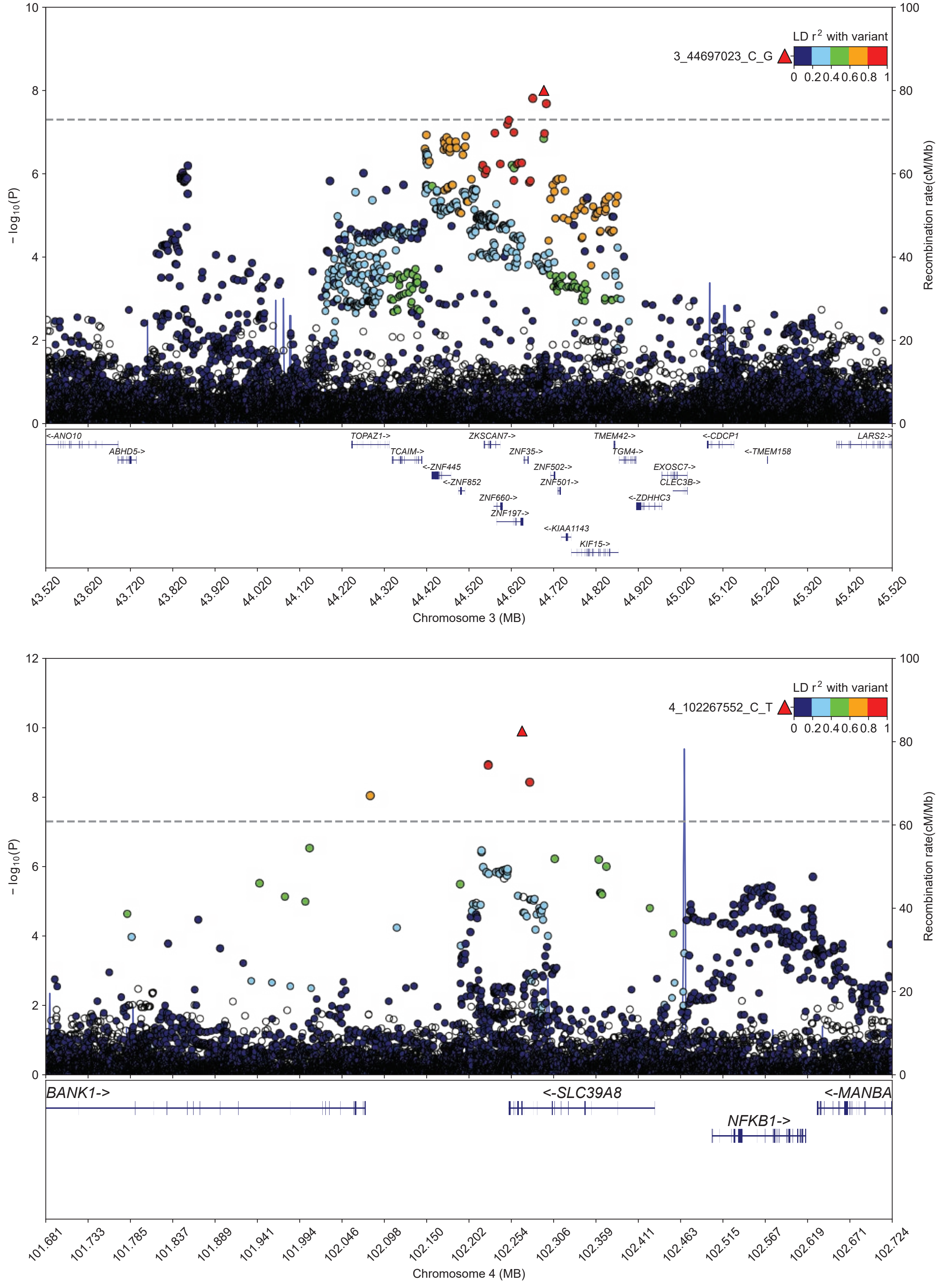

# S3

c

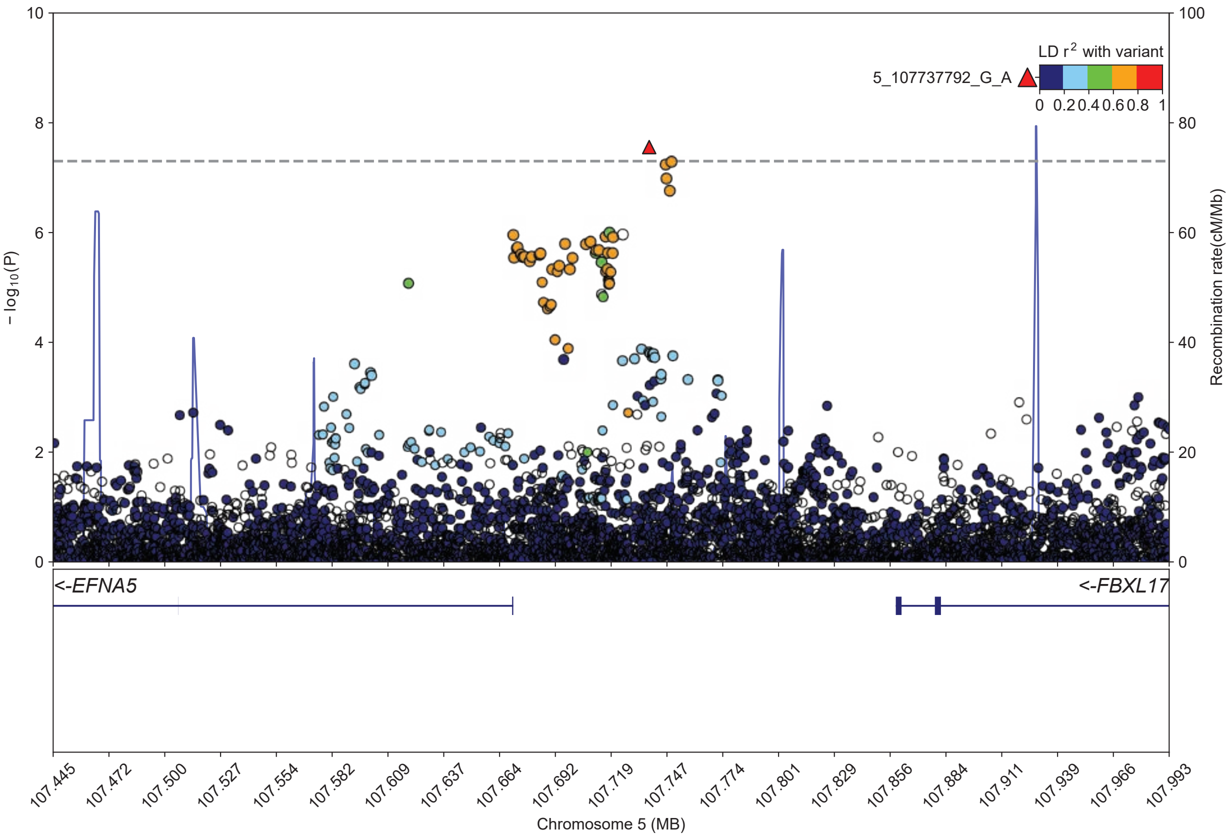
LD r2 with variant
5_107737792_G_A
0 0.2 0.4 0.6 0.8 1
− log10(P)
Recombination rate(cM/Mb)
<-EFNA5
<-FBXL17
107.445
107.472
107.500
107.527
107.554
107.582
107.609
107.637
107.664
107.692
107.719
107.747
107.774
107.801
107.829
107.856
107.884
107.911
107.939
107.966
107.993
Chromosome 5 (MB)

d

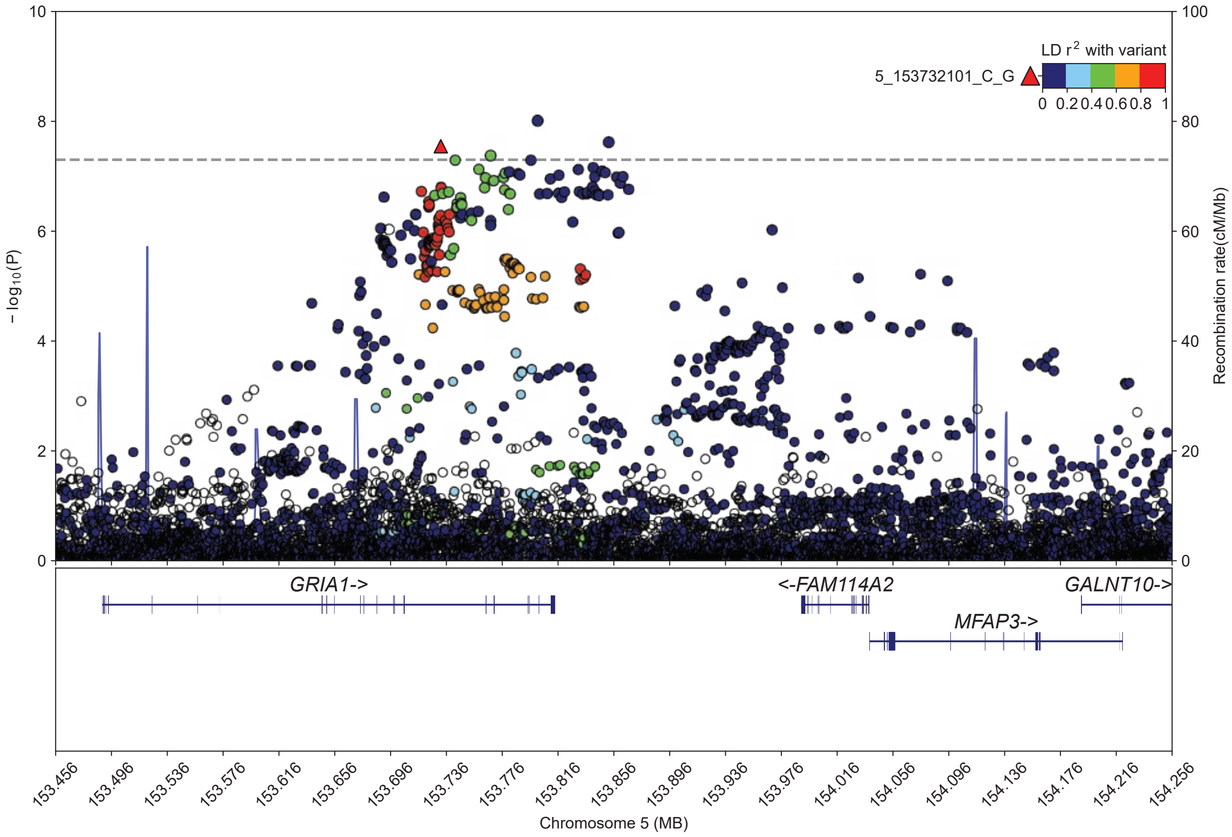
LD r2 with variant
5_153732101_C_G
0 0.2 0.4 0.6 0.8 1
− log10(P)
Recombination rate(cM/Mb)
GRIA1->
<-FAM114A2
GALNT10->
MFAP3->
153.456
153.496
153.536
153.576
153.616
153.656
153.696
153.736
153.776
153.816
153.856
153.896
153.936
153.976
154.016
154.056
154.096
154.136
154.176
154.216
154.256
Chromosome 5 (MB)

# S3

**e**

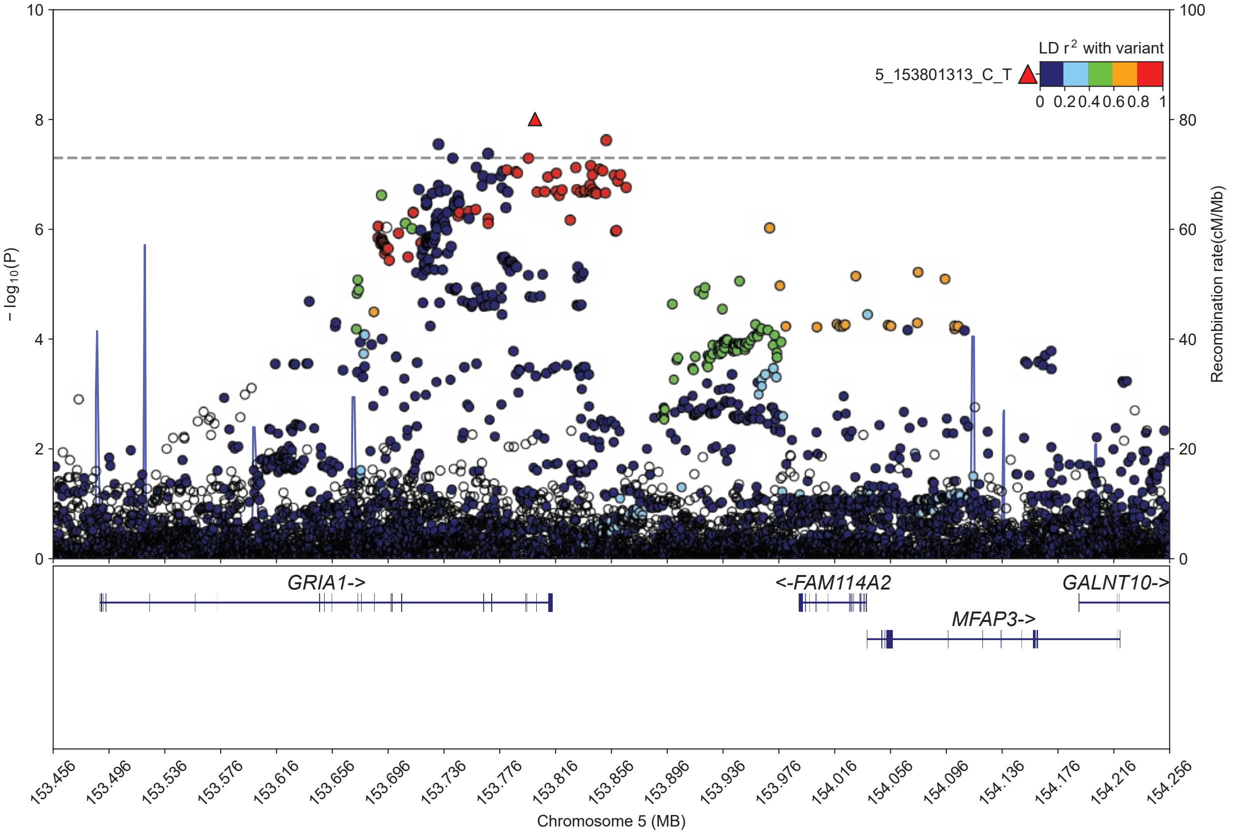

LD r2 with variant
5_153801313_C_T
0 0.2 0.4 0.6 0.8 1
− log10(P)
Recombination rate(cM/Mb)
GRIA1->
<-FAM114A2
GALNT10->
MFAP3->
153.456 153.496 153.536 153.576 153.616 153.656 153.696 153.736 153.776 153.816 153.856 153.896 153.936 153.976 154.016 154.056 154.096 154.136 154.176 154.216 154.256
Chromosome 5 (MB)


**f**

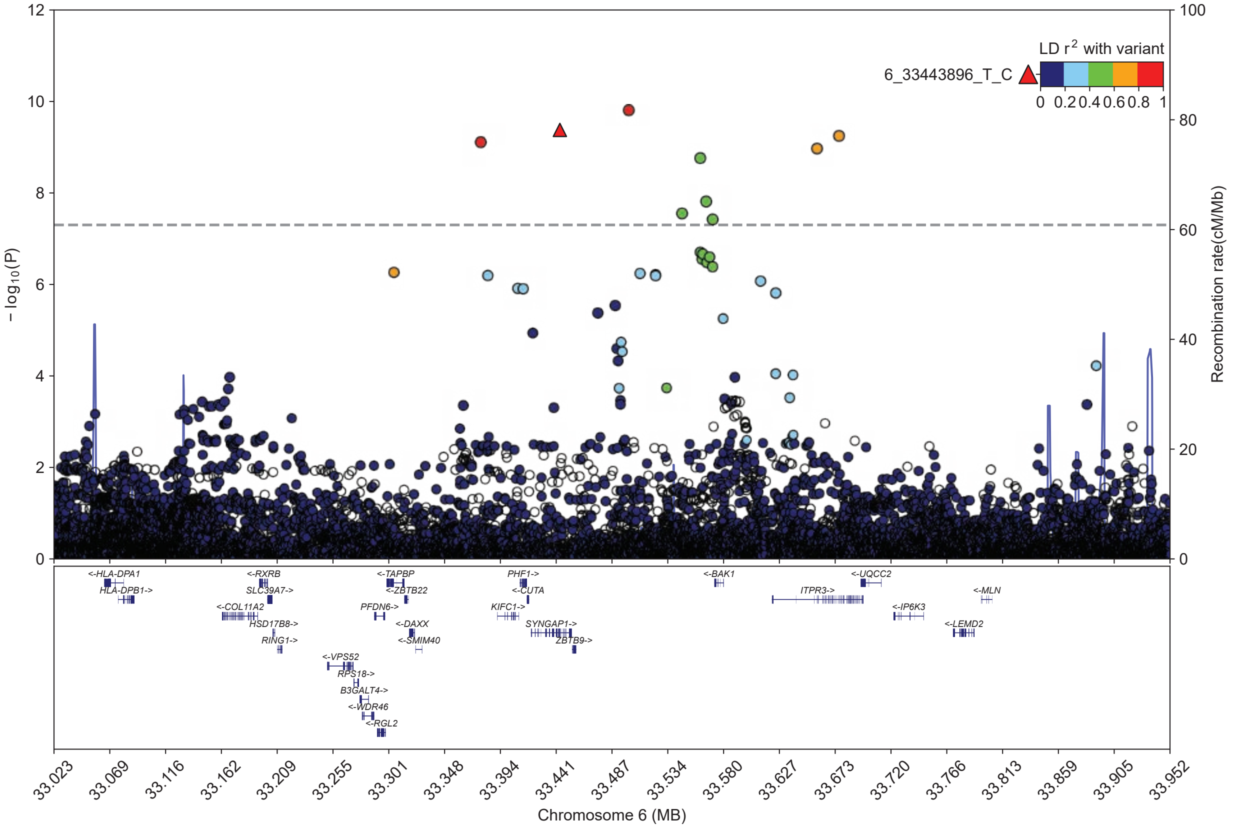

LD r2 with variant
6_33443896_T_C
0 0.2 0.4 0.6 0.8 1
− log10(P)
Recombination rate(cM/Mb)
<-HLA-DPA1
HLA-DPB1->
<-RXRB
SLC39A7->
<-COL11A2
HSD17B8->
RING1->
<-VPS52
RPS18->
B3GALT4->
<-WDR46
<-RGL2
<-TAPBP
<-ZBTB22
PFDN6->
<-DAXX
<-SMIM40
PHF1->
<-CUTA
KIFC1->
SYNGAP1->
ZBTB9->
<-BAK1
ITPR3->
<-UQCC2
<-IP6K3
<-MLN
<-LEMD2
33.023 33.069 33.116 33.162 33.209 33.255 33.301 33.348 33.394 33.441 33.487 33.534 33.580 33.627 33.673 33.720 33.766 33.813 33.859 33.905 33.952
Chromosome 6 (MB)

# S3

g

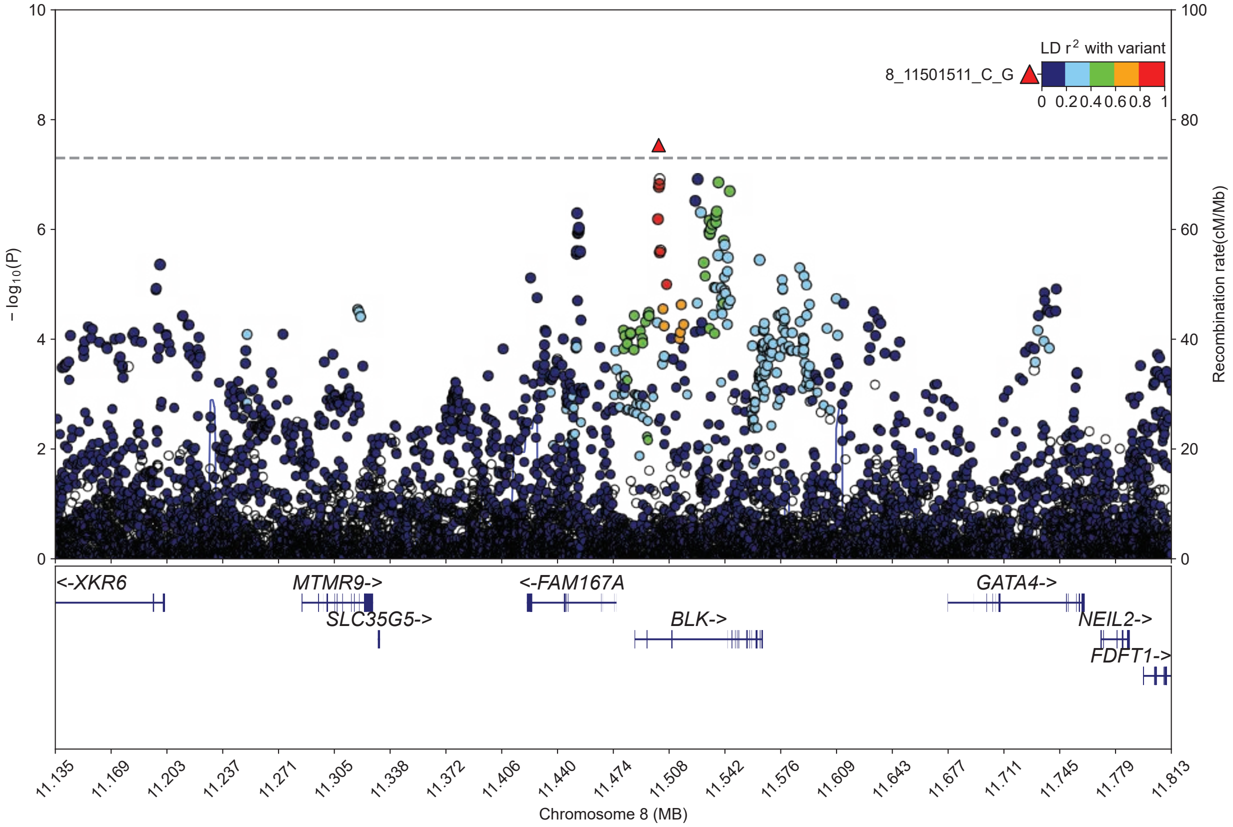
LD r2 with variant
8_11501511_C_G
0 0.2 0.4 0.6 0.8 1
− log10(P)
Recombination rate(cM/Mb)
<-XKR6
MTMR9->
SLC35G5->
<-FAM167A
BLK->
GATA4->
NEIL2->
FDFT1->
Chromosome 8 (MB)

h

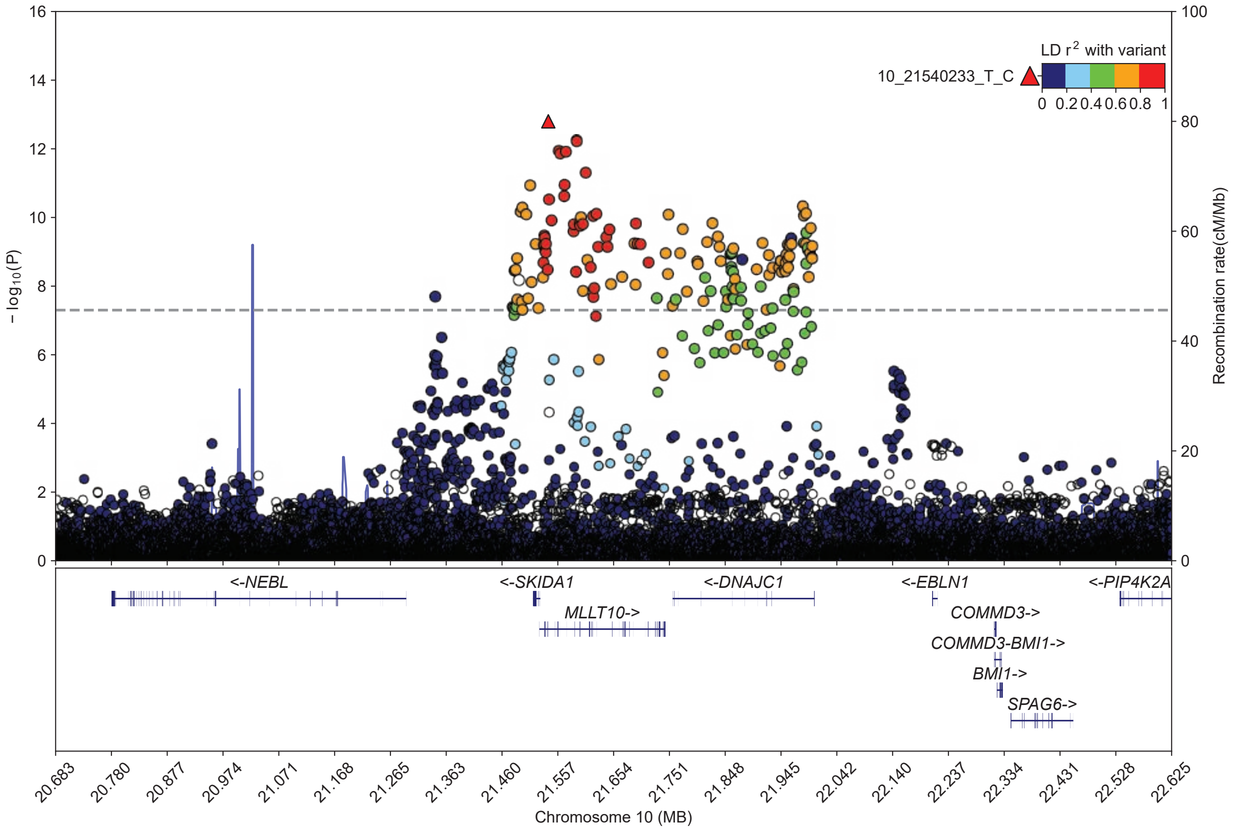
LD r2 with variant
10_21540233_T_C
0 0.2 0.4 0.6 0.8 1
− log10(P)
Recombination rate(cM/Mb)
<-NEBL
<-SKIDA1
MLLT10->
<-DNAJC1
<-EBLN1
COMMD3->
COMMD3-BMI1->
BMI1->
SPAG6->
<-PIP4K2A
Chromosome 10 (MB)

# S3

i

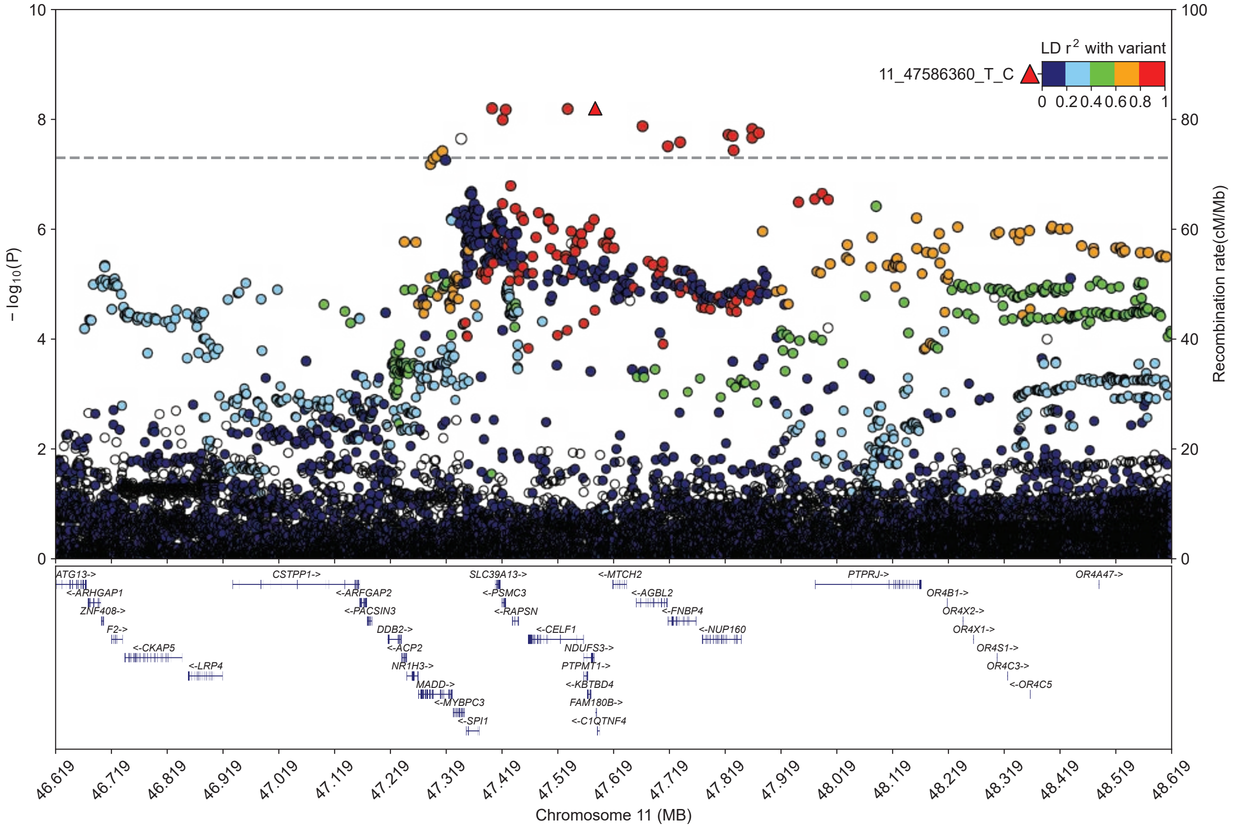

LD r2 with variant
11_47586360_T_C
0 0.2 0.4 0.6 0.8 1
− log10(P)
Recombination rate(cM/Mb)
0
2
4
6
8
10
0
20
40
60
80
100
ATG13->
<-ARHGAP1
ZNF408->
F2->
<-CKAP5
<-LRP4
CSTPP1->
<-ARFGAP2
<-PACSIN3
DDB2->
<-ACP2
NR1H3->
MADD->
<-MYBPC3
<-SPI1
SLC39A13->
<-PSMC3
<-RAPSN
<-CELF1
NDUFS3->
PTPMT1->
<-KBTBD4
FAM180B->
<-C1QTNF4
<-MTCH2
<-AGBL2
<-FNBP4
<-NUP160
PTPRJ->
OR4B1->
OR4X2->
OR4X1->
OR4S1->
OR4C3->
<-OR4C5
OR4A47->
46.619
46.719
46.819
46.919
47.019
47.119
47.219
47.319
47.419
47.519
47.619
47.719
47.819
47.919
48.019
48.119
48.219
48.319
48.419
48.519
48.619
Chromosome 11 (MB)


j

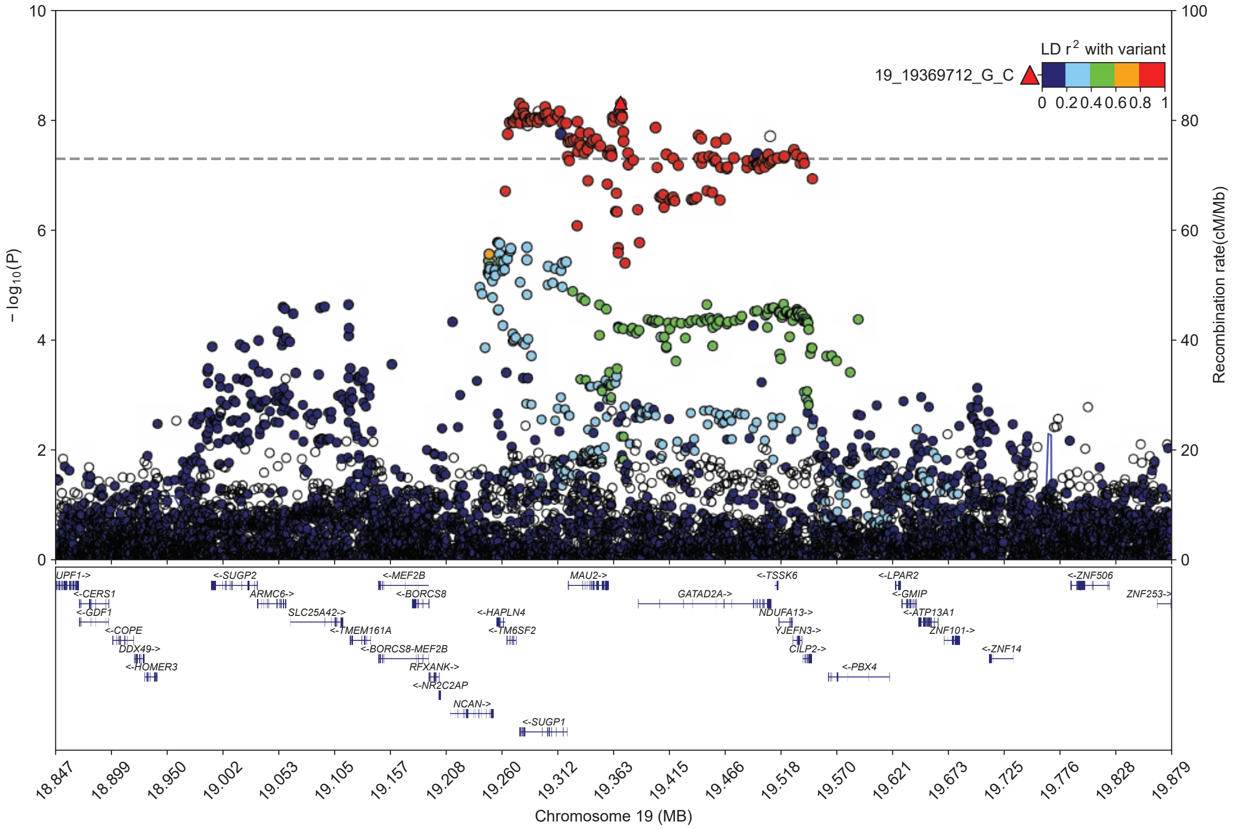

LD r2 with variant
19_19369712_G_C
0 0.2 0.4 0.6 0.8 1
− log10(P)
Recombination rate(cM/Mb)
0
2
4
6
8
10
0
20
40
60
80
100
UPF1->
<-CERS1
<-GDF1
<-COPE
DDX49->
<-HOMER3
<-SUGP2
ARMC6->
SLC25A42->
<-TMEM161A
<-BORCS8-MEF2B
<-MEF2B
<-BORCS8
RFXANK->
<-NR2C2AP
NCAN->
<-HAPLN4
<-TM6SF2
<-SUGP1
MAU2->
GATAD2A->
<-TSSK6
NDUFA13->
YJEFN3->
CILP2->
<-PBX4
<-LPAR2
<-GMIP
<-ATP13A1
ZNF101->
<-ZNF14
<-ZNF506
ZNF253->
18.847
18.899
18.950
19.002
19.053
19.105
19.157
19.208
19.260
19.312
19.363
19.415
19.466
19.518
19.570
19.621
19.673
19.725
19.776
19.828
19.879
Chromosome 19 (MB)

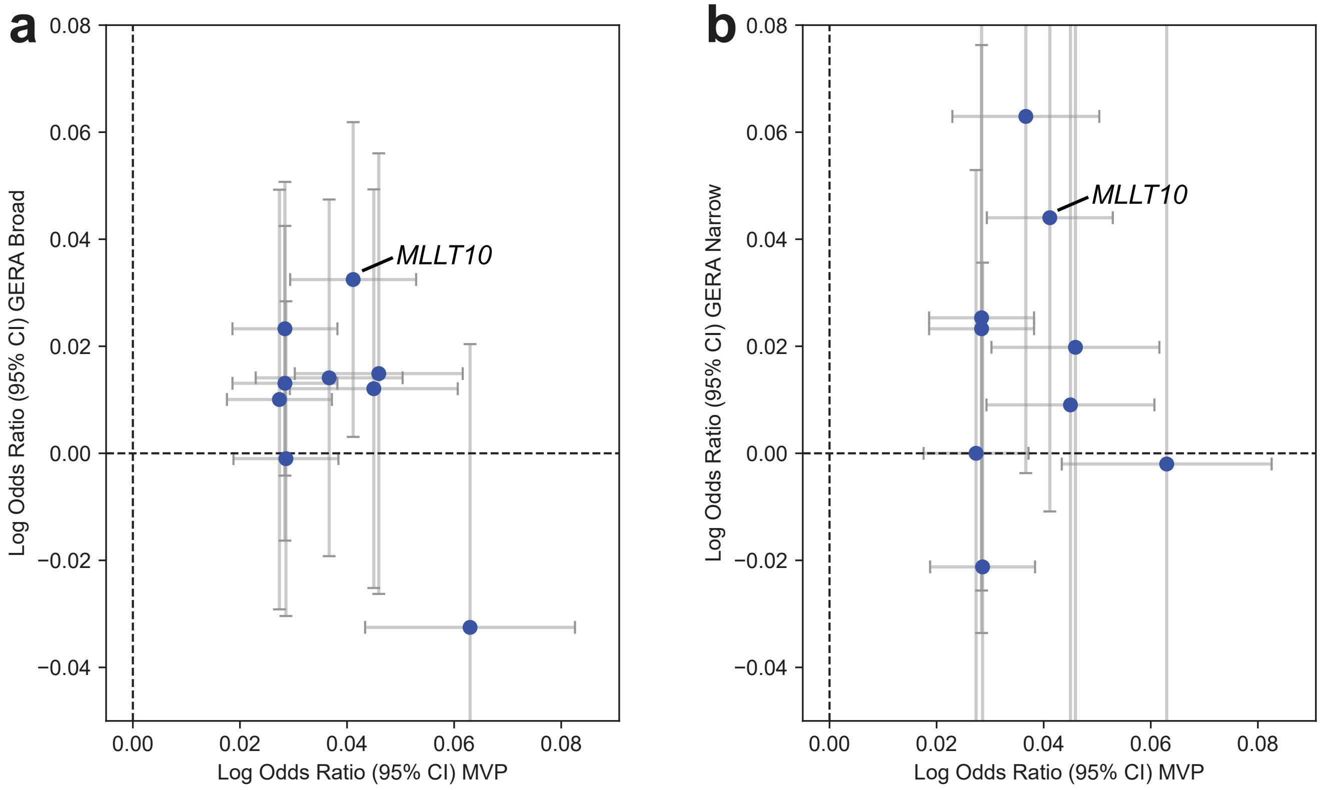


**Figure S4. Comparison of effect sizes at GWAS loci between MVP and the GERA replication cohorts. a)** MVP multi-ancestry meta-analysis (132,637 cases and 352,201 controls) versus the GERA *broad* phenotype (16,025 cases and 54,818 controls). **b)** MVP multi-ancestry meta-analysis versus the stricter GERA *narrow* phenotype (3,317 cases and 54,516 controls). Log odds ratios with 95% confidence intervals are shown. One GWAS locus (rs191549504, at *SYNGAP1*) was not genotyped in GERA. The *MLLT10* locus (rs12779865) is labeled.

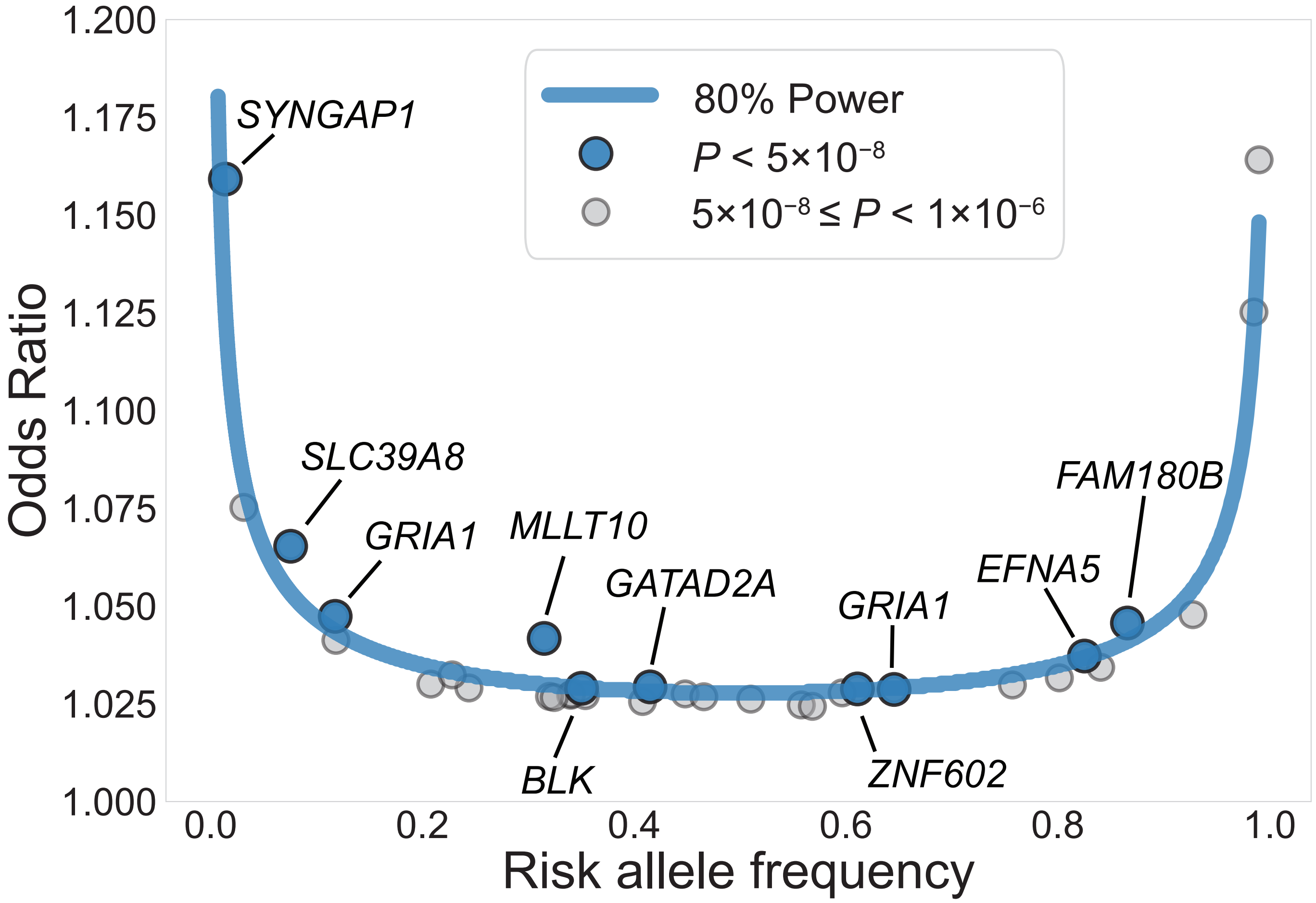


**Figure S5. Comparison of effect size and allele frequency for GWAS loci.** Scatter plot of odds ratio versus risk allele frequency for risk loci in the multi-ancestry meta-analysis (132,637 cases and 352,201 controls). The blue line denotes 80% power to achieve genome-wide significance. Independent genome-wide significant loci are highlighted in blue and labeled by nearest gene. Additional independent loci reaching a suggestive significance threshold ($P < 1\times10^{-6}$) are shown in gray.

**VA Million Veteran Program:**
**Core Acknowledgements for Publications**
**May 2024**

### MVP Program Office

- Sumitra Muralidhar, Ph.D., Program Director
  US Department of Veterans Affairs, 810 Vermont Avenue NW, Washington, DC 20420
- Jennifer Moser, Ph.D., Associate Director, Scientific Programs
  US Department of Veterans Affairs, 810 Vermont Avenue NW, Washington, DC 20420
- Jennifer E. Deen, B.S., Associate Director, Cohort & Public Relations
  US Department of Veterans Affairs, 810 Vermont Avenue NW, Washington, DC 20420

### MVP Executive Committee

- Co-Chair: Philip S. Tsao, Ph.D.
  VA Palo Alto Health Care System, 3801 Miranda Avenue, Palo Alto, CA 94304
- Co-Chair: Sumitra Muralidhar, Ph.D.
  US Department of Veterans Affairs, 810 Vermont Avenue NW, Washington, DC 20420
- J. Michael Gaziano, M.D., M.P.H.
  VA Boston Healthcare System, 150 S. Huntington Avenue, Boston, MA 02130
- Elizabeth Hauser, Ph.D.
  Durham VA Medical Center, 508 Fulton Street, Durham, NC 27705
- Amy Kilbourne, Ph.D., M.P.H.
  VA HSR&D, 2215 Fuller Road, Ann Arbor, MI 48105
- Michael Matheny, M.D., M.S., M.P.H.
  VA Tennessee Valley Healthcare System, 1310 24th Ave. South, Nashville, TN 37212
- Dave Oslin, M.D.
  Philadelphia VA Medical Center, 3900 Woodland Avenue, Philadelphia, PA 19104
- Deepak Voora, MD
  Durham VA Medical Center, 508 Fulton Street, Durham, NC 27705

### MVP Co-Principal Investigators

- J. Michael Gaziano, M.D., M.P.H.
  VA Boston Healthcare System, 150 S. Huntington Avenue, Boston, MA 02130
- Philip S. Tsao, Ph.D.
  VA Palo Alto Health Care System, 3801 Miranda Avenue, Palo Alto, CA 94304

### MVP Core Operations

- Jessica V. Brewer, M.P.H., Director, MVP Cohort Operations
  VA Boston Healthcare System, 150 S. Huntington Avenue, Boston, MA 02130
- Mary T. Brophy M.D., M.P.H., Director, VA Central Biorepository
  VA Boston Healthcare System, 150 S. Huntington Avenue, Boston, MA 02130
- Kelly Cho, M.P.H, Ph.D., Director, MVP Phenomics

VA Boston Healthcare System, 150 S. Huntington Avenue, Boston, MA 02130

- Lori Churby, B.S., Director, MVP Regulatory Affairs
  VA Palo Alto Health Care System, 3801 Miranda Avenue, Palo Alto, CA 94304
- Scott L. DuVall, Ph.D., Director, VA Informatics and Computing Infrastructure (VINCI)
  VA Salt Lake City Health Care System, 500 Foothill Drive, Salt Lake City, UT 84148
- Saiju Pyarajan Ph.D., Director, Data and Computational Sciences
  VA Boston Healthcare System, 150 S. Huntington Avenue, Boston, MA 02130
- Robert Ringer, Pharm.D., Director, VA Albuquerque Central Biorepository
  New Mexico VA Health Care System, 1501 San Pedro Drive SE, Albuquerque, NM 87108
- Luis E. Selva, Ph.D., Director, MVP Biorepository Coordination
  VA Boston Healthcare System, 150 S. Huntington Avenue, Boston, MA 02130
- Shahpoor (Alex) Shayan, M.S., Director, MVP PRE Informatics
  VA Boston Healthcare System, 150 S. Huntington Avenue, Boston, MA 02130
- Brady Stephens, M.S., Principal Investigator, MVP Information Center
  Canandaigua VA Medical Center, 400 Fort Hill Avenue, Canandaigua, NY 14424
- Stacey B. Whitbourne, Ph.D., Director, MVP Cohort Development and Management
  VA Boston Healthcare System, 150 S. Huntington Avenue, Boston, MA 02130

**MVP Publications and Presentations Committee**

- Co-Chair: Themistocles L. Assimes, M.D., Ph. D
  VA Palo Alto Health Care System, 3801 Miranda Avenue, Palo Alto, CA 94304
- Co-Chair: Adriana Hung, M.D.; M.P.H
  VA Tennessee Valley Healthcare System, 1310 24th Ave. South, Nashville, TN 37212
- Co-Chair: Henry Kranzler, M.D.
  Philadelphia VA Medical Center, 3900 Woodland Avenue, Philadelphia, PA 19104
